# Clinical presentation, disease course and outcome of COVID-19 in hospitalized patients with and without pre-existing cardiac disease – a cohort study across eighteen countries

**DOI:** 10.1101/2021.03.11.21253106

**Authors:** CAPACITY-COVID collaborative consortium and LEOSS Study Group, M Linschoten, A Uijl, A Schut, CEM Jakob, LR Romão, RM Bell, E McFarlane, M Stecher, AGM Zondag, EPA van Iperen, W Hermans-van Ast, NC Lea, J Schaap, LS Jewbali, PC Smits, RS Patel, A Aujayeb, DP Ripley, M Saxena, C Spinner, GP McCann, AJ Moss, E Parker, S Borgmann, E Tessitore, S Rieg, MT Kearney, R Byrom-Goulthorp, M Hower, AK Al-Ali, AM Alshehri, AN Alnafie, M Alshahrani, YA Almubarak, FA Al-Muhanna, AM Al-Rubaish, F Hanses, AC Shore, C Ball, CM Anning, MM Rüthrich, PR Nierop, MJGT Vehreschild, SRB Heymans, MTHM Henkens, AG Raafs, ICC van der Horst, BCT van Bussel, FJH Magdelijns, J Lanznaster, PY Kopylov, OV Blagova, K Wille, YM Pinto, JA Offerhaus, H Bleijendaal, C Piepel, JM ten Berg, WL Bor, M Maarse, C Römmele, RA Tio, NH Sturkenboom, L Tometten, CA den Uil, NTB Scholte, AL Groenendijk, S Dolff, LE Zijlstra, AD Hilt, M von Bergwelt-Baildon, BE Groenemeijer, U Merle, PM van der Zee, EA van Beek, K Rothfuss, FVY Tjong, ACJ van der Lingen, MZH Kolk, N Isberner, PS Monraats, M Magro, WRM Hermans, M Kochanek, G Captur, RJ Thomson, S Nadalin, GCM Linssen, T Veneman, R Zaal, C Degenhardt, FMAC Martens, EA Badings, R Strauss, AG Zaman, M Alkhalil, S Prasad, B Grüner, HE Haerkens-Arends, L Eberwein, P Dark, D Lomas, J vom Dahl, DO Verschure, K Hellwig, A Mosterd, D Rauschning, DJ van der Heijden, M Neufang, M van Hessen, C Raichle, L Montagna, SG Mazzilli, M Bianco, T Westhoff, A Shafiee, B Hedayat, E Saneei, H Porhosseini, B Jensen, L Gabriel, AG Er, BLJH Kietselaer, J Schubert, P Timmermans, P Messiaen, A Friedrichs, FS van den Brink, P Woudstra, J Trauth, MIA Ribeiro, K de With, MMJM van der Linden, JT Kielstein, R Macías Ruiz, W Guggemos, E Hellou, P Markart, HAM van Kesteren, D Heigener, JK de Vries, S Stieglitz, JB Baltazar, I Voigt, DJ van de Watering, M Milovanovic, J Redón, MJ Forner, J Rüddel, KW Wu, J Nattermann, LI Veldhuis, ICD Westendorp, C Riedel, JM Kwakkel-van Erp, S van Ierssel, EM van Craenenbroeck, L Walter, J de Sutter, M Worm, JT Drost, A Moriarty, R Salah, N Charlotte, AJM van Boxem, HGR Dorman, AC Reidinga, P van der Meer, E Wierda, HPAA van Veen, CE Delsing, MFL Meijs, RMA van de Wal, C Weytjens, RS Hermanides, ME Emans, NYY al-Windy, AMH Koning, DAAM Schellings, RL Anthonio, C Bucciarelli-Ducci, M Caputo, PHM Westendorp, AFM Kuijper, CEE van Ofwegen-Hanekamp, AM Persoon, J Seelig, P van der Harst, HJ Siebelink, M van Smeden, S Williams, L Pilgram, WH van Gilst, RG Tieleman, B Williams, FW Asselbergs

## Abstract

**Aims:** Patients with cardiac disease are considered high risk for poor outcomes following hospitalization with COVID-19. The primary aim of this study was to evaluate heterogeneity in associations between various heart disease subtypes and in-hospital mortality.

**Method and results:** We used data from the CAPACITY-COVID registry and LEOSS study. Multivariable Poisson regression models were fitted to assess the association between different types of pre-existent heart disease and in-hospital mortality. 16,511 patients with COVID-19 were included (21.1% aged 66 – 75 years; 40.2% female) and 31.5% had a history of heart disease. Patients with heart disease were older, predominantly male and often had other comorbid conditions when compared to those without. Mortality was higher in patients with cardiac disease (29.7%; n=1545 versus 15.9%; n=1797). However, following multivariable adjustment this difference was not significant (adjusted risk ratio (aRR) 1.08 [95% CI 1.02 – 1.15; p-value 0.12 (corrected for multiple testing)]). Associations with in-hospital mortality by heart disease subtypes differed considerably, with the strongest association for heart failure aRR (1.19 [1.10 – 1.30]; p-value <0.018) particularly for severe NYHA III/IV) heart failure (aRR 1.41 [95% CI 1.20 – 1.64; p-value <0.018]. None of the other heart disease subtypes, including ischemic heart disease, remained significant after multivariable adjustment. Serious cardiac complications were diagnosed in <1% of patients.

**Conclusion:** Considerable heterogeneity exists in the strength of association between heart disease subtypes and in-hospital mortality. Of all patients with heart disease, those with heart failure are at greatest risk of death when hospitalized with COVID-19. Serious cardiac complications are rare.

## Introduction

Coronavirus disease 2019 (COVID-19) has rapidly spread across the globe since December 2019, leading to more than 175 million confirmed cases and 3.5 million fatalities as of the 18^th^ of June 2021.^1^ Although the disease is not as lethal (case fatality ratio (CFR) ∼0.3 – 1%)^2,3^ as the Middle-East respiratory syndrome (MERS; CFR ∼35%)^4^ and the severe acute respiratory syndrome (SARS; CFR 14 – 15%)^5^, it has become clear that morbidity and mortality is much higher than pandemic influenza (CFR 0.1%)^6,7^, especially among the elderly^3^.

Studies show that a significant number of patients who develop severe symptoms of COVID-19 have underlying comorbidities, of which cardiovascular disease (CVD) is reported in 10 – 30% of inpatients in Western-European and American cohorts.^8-11^ Patients with pre-existing cardiac disease have consistently been reported to be at increased risk of an unfavorable outcome both among the general population and those requiring hospitalization when compared to patient without these conditions.^8,12^ In one of the largest cohort studies of hospitalized patients thus far (n=20,133), chronic cardiac disease was significantly associated with mortality (adjusted HR 1.16; 95% confidence interval (CI) 1.08 – 1.24).^8^ Another study by Fried et al. (n=11,721) across 38 states in the United States, found an adjusted odds ratio (aOR) of 1.22 (95% CI 1.06 – 1.41) and 1.44 (95% CI 1.27 – 1.63) for mechanical ventilation and death respectively related to cardiac disease.^11^ The Chinese Center for Disease Control report a CFR five times higher among those with CVD compared to patients without any comorbidities.^13^ These observations are in line with previous studies among patients with influenza and other respiratory tract infections.^14-15^

Previous studies have predominantly evaluated the association between having any chronic cardiac disease and COVID-19 related mortality, where all cardiac disease subtypes are analyzed together.^8,11,12^ However, from a clinical point of view, it is likely that not all cardiac diseases mediate a similar risk. Increasing our understanding of the clinical course of COVID-19 in patients across different heart disease subtypes is of pivotal importance. Firstly, it can provide guidance for health care professionals in the management of these patients and would better inform shielding guidelines. Secondly, it can bring some clarity for patients, concerned about how their own cardiac disease influences their risk from COVID-19.^16^ Reducing these concerns might also have a positive impact on health care seeking behavior during the pandemic, diminishing the detrimental collateral damage the outbreak has provoked in patients with heart disease who are afraid of attending hospitals.^17^

The aim of the current study was to investigate whether there is heterogeneity in the strength of association per heart disease subtype and in-hospital mortality. Furthermore, we describe the disease trajectory of COVID-19 in hospitalized patients with- and without pre-existing cardiac disease, from documentation at hospital admission to discharge or death, including the prevalence of cardiac complications.

## Methods

### Study design and setting

For this study we used data collected in the CAPACITY-COVID registry (www.capacity-covid.eu) and the Lean Open Survey on SARS-CoV-2 infected patients (LEOSS) study (www.LEOSS.net).

CAPACITY-COVID is a multinational patient registry specifically established to determine the role of CVD in the COVID-19 pandemic (NCT04325412).^18^ All adult patients (≥18 years) hospitalized with confirmed or highly suspected COVID-19 are eligible for inclusion in the registry. The extent and scope of inclusion varies per site, depending on local resources and preference. A majority of participating centers (n=56) use a non-selective inclusion, i.e., every adult patient with (highly suspected) COVID-19 or a random sample is included in the registry, and 18 centers applied a selective inclusion of only patients for whom a cardiologist has been consulted, only patients with a history of CVD or cardiovascular risk factors or a selection based on department of admission i.e., only patients admitted to the ward or intensive care unit (ICU). Since the launch of the registry in March 2020, 74 centers across 13 countries have joined the consortium.

Within CAPACITY-COVID, the ISARIC core case report form^19^ has been used as the core data set which was extended with ∼400 additional variables to capture in-depth information regarding cardiovascular history, the use of cardiovascular medications, cardiac investigations such as ECG and echocardiography and cardiovascular outcomes. The data dictionary is available online (www.capacity-covid.eu). Only data generated during routine clinical care is collected and patients do not undergo any additional investigations for the purpose of this registry. Data is collected in a REDCap database after pseudonymization which is managed by the University Medical Center Utrecht, Utrecht, the Netherlands.

Variable definitions handled in CAPACITY-COVID are incorporated in the REDCap Case Report Form (CRF) and can be found online among the study documents at: https://capacity-covid.eu/for-professionals/. For the registration of cardiac complication, the diagnostic criteria of the European Society of Cardiology (ESC) guidelines for myocarditis^20^, pericarditis^21^ endocarditis^22^ and acute coronary syndrome^23^ were incorporated to minimize heterogeneity in the adjudication of these events. For arrhythmia’s, the ACC/AHA/HRS 2006 key data elements and definitions for electrophysiological studies and procedures was used.^24^ In absence of a definition, a clinical diagnosis as indicated in the Electronic Health Record (EHR) was handled.

LEOSS is a European multicenter cohort study established in March 2020, collecting information on both hospitalized and ambulant patients with laboratory confirmed COVID-19. A detailed description of the study design has been previously reported.^25^ In short, patients of all ages can be included in LEOSS. Case collection is anonymized, realized by amongst other the absence of any variables containing directly identifying information in the case report form, only a small subset of variables associated with a high risk of re-identification and the categorized collection of continuous variables (such as age).^26^ The CRF of LEOSS can be provided upon request. Currently centers from Austria, Belgium, Bosnia and Herzegovina, Germany, Italy, Latvia Spain, Switzerland, Turkey and the United Kingdom contribute data to the registry. The data collection is coordinated by the University Hospital of Cologne in Germany.

### Study population

We excluded patients only treated in an ambulatory setting, children (age <18 years) and patients for which the region of inclusion, COVID-19 status, admission date, history of cardiac disease, age, sex or outcome were unconfirmed. All hospitalized patients aged ≥18 years with a laboratory confirmed SARS-CoV-2 infection registered between March and May 2021 were included. A list of all participating sites that contributed with data to the current study is provided in **Table S1**. The informed consent procedure varied per study site, following local and national rules and regulations during the pandemic. Within CAPACITY-COVID, a majority of participating sites handled an opt-out approach, where patients received written information during or after hospital admission. For sites in the UK, informed consent was not required under emergency legislation during the pandemic. Inclusion into LEOSS did not require informed consent due to anonymous case collection. Medical ethics approval was obtained nationally or independently for each participating site complying with the Declaration of Helsinki.

### Statistical analysis

Continuous variables in CAPACITY-COVID were categorized to align with LEOSS prior to merging the datasets from these two different sources. Multiple imputation (R package *mice*) was performed to deal with missingness across baseline variables required for the regression models through the generation of 10 imputed datasets. The following variables were included in the multiple imputation model: all variables indicated with an asterisk in **Table 1-3**, and furthermore presence of pre-existing cardiac disease, arrhythmia/conduction disorder, heart failure, coronary artery disease (CAD), valvular heart disease, month and year of hospital admission, extent of inclusion (all/random sample vs. selected inclusion as described above), cohort (CAPACITY-COVID vs. LEOSS) and region of inclusion (Central Europe, Netherlands/Belgium, Middle – East, Southern-Europe and the United Kingdom). Baseline variables with >40% missingness were excluded from further analysis.

For the main analyses, multivariable modified Poisson models with robust standard errors were used to estimate the association between a history of cardiac disease and in-hospital mortality across the pooled ten imputed datasets. Heterogeneity in associations was also determined across various clinically relevant subgroups including age (≤65 and >65), sex, body mass index (BMI) (<30 and ≥30), diabetes, hypertension, chronic kidney disease (CKD) and chronic obstructive pulmonary disease (COPD). In the secondary analyses, associations between pre-defined specific types of heart disease and the in-hospital mortality were determined. The following heart disease subtypes were analyzed: arrhythmias/conduction disorders, CAD, myocardial infarction (MI), heart failure and valvular heart disease. To be able to determine differences in the association between the pre-defined cardiac disease subtypes and in-hospital mortality in adults versus the elderly, analyses were also performed in patients ≤65 years and >65 years. The cut-off was set at 65 since most COVID-19 vaccination strategies in Europe have defined the elderly as those >65 years of age.^27^

All analyses were adjusted for the following covariates: age, sex, BMI, diabetes, hypertension, CKD, COPD and geographic region of inclusion (see above). For the sensitivity analyses we excluded patients included by centers handling a selective inclusion strategy as previously outlined. In addition, we assessed the direction and magnitude of the associations based on registry of inclusion (CAPACITY vs. LEOSS), next to fitting models in the dataset of the combined cohorts.

Logistic regression on the non-imputed dataset was used to identify association of the most prevalent COVID-19 symptoms at presentation and eight different age categories in the total cohort and after stratification on pre-existent cardiac disease. Due to low numbers of young patients with a history of cardiac disease, the four lowest age categories were merged in the stratified analyses (18 – 55 years).

Results of the modified Poisson regression models are reported as risk ratios (RR) with 95% confidence intervals (CI)) and the logistic regression models as odds ratios (ORs) with 95% CI’s. Statistical significance was set at an alpha of 5% and all hypothesis tests were two-sided. The regression analyses were corrected for multiple testing using the Holm-Bonferroni method. Continuous variables were summarized as means (SD) or medians [IQR] and categorical variables as counts (%). All analyses were performed in R Studio (version 1.3.959, Vienna, Austria).

## Results

### Baseline characteristics

In total, 20,954 patients had been included in CAPACITY-COVID or LEOSS between March and May 2021. After applying exclusion criteria, 16,511 patients from eighteen countries were retained for the final analysis (**Figure S1**). Baseline characteristics stratified by the presence of pre-existing cardiac disease and age are summarized in **Table 1**.

Most patients were aged between 76 – 85 (n=3,720; 22.5%), and more than half the cohort was composed of individuals >65 years (n=8,861; 53.7%). Most patients were white (n=12,120; 84.5%), predominantly male (n=9,864; 59.8%), and 68.0% (n=7,080 had a BMI >25 kg/m^2^. At baseline, almost one third of patients (n=5,198; 31.5%) had pre-existing heart disease of which cardiac arrhythmias/conduction disorders (n=2,503; 15.3%) and CAD (n=2,420; 14.8%) were most common (**Table S2**). In total, 1,314 patients (8.1%) had been previously diagnosed with heart failure. Other frequent major comorbidities were diabetes (n=4,031; 24.9%) and CKD (n=2,196; 13.5%).

Compared to patients without a history of cardiac disease, patients with pre-existing heart disease were generally older, more often male (63.6% vs. 58.1%) and had a higher burden of cardiovascular risk factors and other comorbid conditions at baseline (**Table 1**). Detailed phenotyping of underlying pre-existing cardiac diseases of patients registered in CAPACITY-COVID, including arrhythmias, conduction disorders, type of ischemic- and valvular heart disease are outlined in **Table S3**. To evaluate heterogeneity across datasets, baseline characteristics were also stratified by the cohort of origin (CAPACITY-COVID vs. LEOSS; **Table S2**). Patients in LEOSS tended to be younger with an overall higher prevalence of heart disease.

### Complaints at admission

The median duration from the onset of symptoms to hospital admission was 6 days [IQR 2 – 9]. Fever, cough and shortness of breath were the most common symptoms at presentation reported in 54.9%, 51.8% and 49.8% of patients respectively (**Table 2**). The odds of having these symptoms varied across age, with fever, cough and dyspnea being reported less frequently in the younger (<45 years) and older (>65 years) age groups (**Figure S2**).

The probability of experiencing a sore throat, anosmia and chest pain declined with age, while fatigue was reported more often with increasing age. Heterogeneity in complaints and vital signs at admission was also assessed by stratification by cohort of inclusion (CAPACITY-COVID vs. LEOSS; **Table S4**). Of symptoms overlapping with CVD, chest pain was most common, reported by 9.2% of patients. Less than five percent of patients experienced (pre) syncope, palpitations, orthopnea or peripheral edema (**Table S4**). After stratification by age, the pattern of complaints did not differ between patients with and without a history of cardiac disease (**Figure S3**).

### Outcomes

The median duration of hospital admission was 9 [5 – 18] days (**Table 3**). More than one in four patients were admitted to a critical care unit (n=3916; 27.6%) with a median length of stay of 12 [6 – 23] days. The proportion of patients admitted to a critical care unit increased with age until 75 years. Patients aged >75 years were predominantly treated on the ward (**Table S5**). Overall, the comorbidity burden among patients on a critical care unit was lower than for patients admitted to the ward only (**Table S5**). Patients that were admitted to a critical care unit tended to be more ill at admission based on vitals and laboratory values at hospital admission (**Table S6**). During hospital admission 20.2% (n=3342) of patients died. Mortality was strongly related to age, with a mortality of 0.8% (n=2) in patients aged 18-25 years and 39.4% (n=652) in patients aged >85 years (**Table S7**). Oxygen saturation levels were lower at admission in those who died, while the levels of inflammatory markers (C-reactive protein and total white blood cell count) were higher (**Table S8**). In patients with cardiac disease 29.7% (n=1545) died during their hospital admission versus 15.9% (n=1797) in patients without chronic heart disease (**Table 3**). In addition to heart disease, other comorbidities were also more prevalent in patients that died during their hospital admission (**Table S7**).

### Cardiac and thromboembolic complications

During hospitalization, serious cardiac complications including myocarditis, MI and new onset heart failure were diagnosed in 0.2% (n=37), 0.6% (n=95) and 1.2% (n=197) of patients respectively (**Table 3**). Other serious cardiac complications registered only in CAPACITY-COVID, including malignant ventricular arrhythmias, endocarditis and pericarditis were also uncommonly diagnosed (<1% of patients; **Table S9**). MI and new onset heart failure was diagnosed more frequently in patients with pre-existing cardiac disease compared to those without (**Table 3**). Among thromboembolic complications, pulmonary embolism was most prevalent being diagnosed in 3.5% (n=569) of patients (**Table 3**). All complications occurred more often in patients admitted to a critical care unit and patients that died in-hospital (**Table S10, S11**), with the difference being most pronounced for pulmonary embolism, which was diagnosed in 10.5% (n=405) the critically ill vs. 1.4 (n=144) among patients only admitted to the ward. Venous thromboembolic complications were diagnosed less frequently among patients with a history of heart disease (**Table 3**).

### Association between prior history of cardiac disease and in-hospital mortality

The multivariable modified Poisson regression model fitted in the total population yielded a non-statistically significant association between any pre-existing cardiac disease and in-hospital mortality (aRR 1.08 [95% CI 1.02 1.15, p-value 0.12])(**Figure 1**). This association was further explored across different clinically relevant subgroups (**Table S12**). Apart from age and sex, displaying a trend towards an interaction with prior heart disease (p-value 0.10 and 0.09 respectively), the subgroup analyses did not reveal any other interactions. Furthermore, sensitivity analyses by the exclusion of patients included from centers that handled a selective inclusion (n=707) did not yield any different results (**Table S13**).

**Figure 1:**
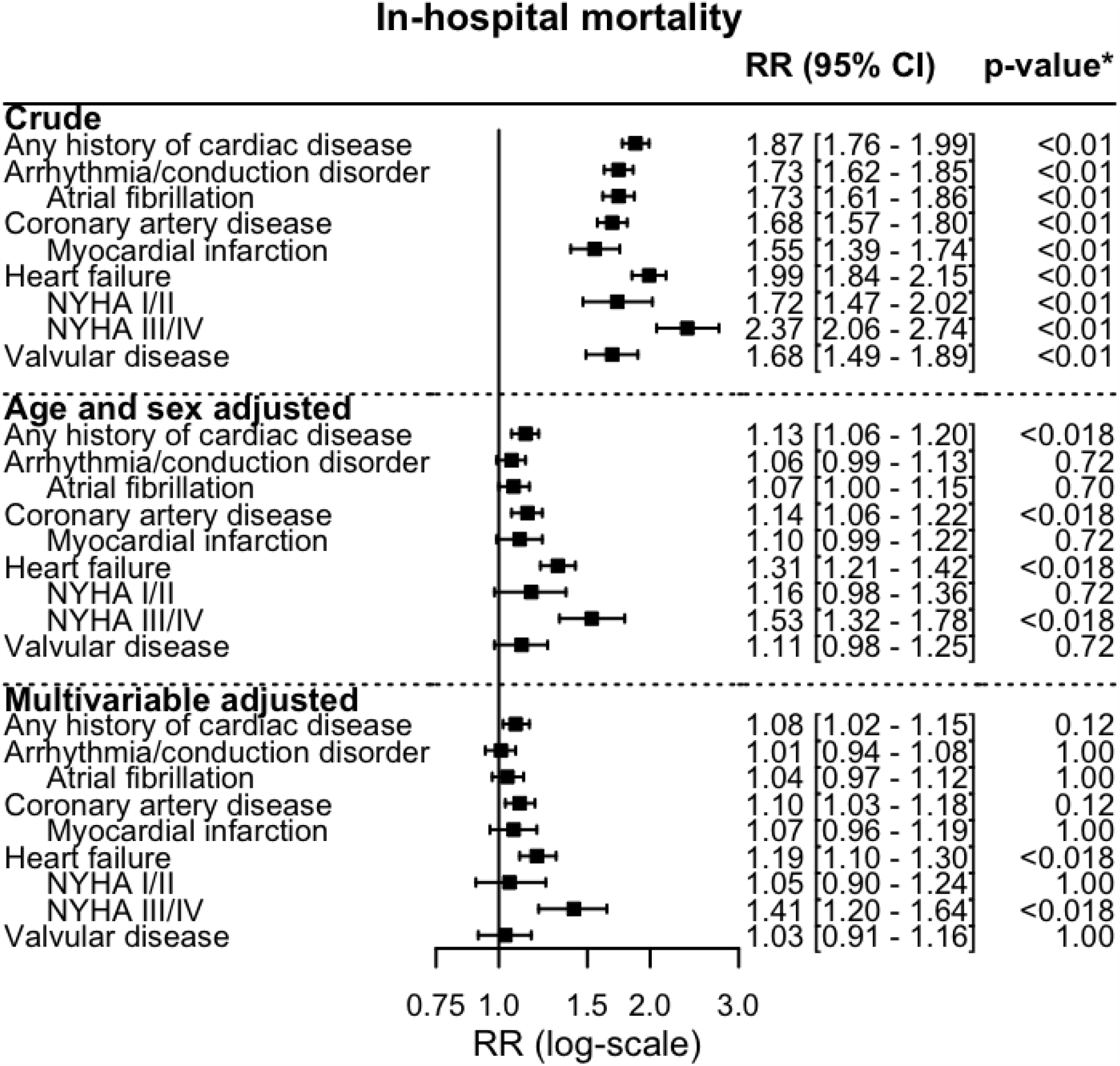
Associations (crude: top; age- and sex adjusted; center, multivariable adjusted; bottom) between any history of cardiac disease, heart disease subtypes and in-hospital mortality. P-values are adjusted for multiple testing.

### Association between different pre-existing cardiac comorbidities and in-hospital mortality

To assess heterogeneity in the associations between different types of heart disease and hospital mortality, modified Poisson regression models were fitted for all pre-specified cardiac disease subgroups. After multivariable adjustment, the strongest association was found for heart failure and in-hospital mortality (aRR 1.19 [95% CI 1.10 – 1.30; p-value <0.018] and in particular for severe (NYHA III/IV) heart failure (aRR 1.41 [95% CI 1.20 – 1.64; p-value <0.018)(**Figure 1**). For the other heart disease subtypes, including ischaemic heart disease, no significant associations with in-hospital mortality were found (**Figure 1, Table S14**).

Since the elderly (>65 years) and adults with comorbidities are defined as two of the main risk groups being prioritized in ongoing vaccination campaigns and statistical interaction was established between pre-existing cardiac disease and age, associations between the various heart disease subtypes and in-hospital mortality were also determined in patients ≤65 years and >65 years (**Figure 2**). However, for none of the heart disease subtypes the interaction with age was found significant. As heterogeneity in treatment intensity may impact the associations between pre-existing heart disease and in-hospitals, we also performed a separate analysis in patients admitted only to the wards vs. patients admitted to a critical care unit. A strong interaction was found between having any history of cardiac disease, arrhythmia/conduction disorders, valvular heart disease and admission to a critical care unit (**Table S15**). The adjusted RR’s between these pre-existing heart conditions and in-hospital mortality overall were lower in patients that had been admitted to a critical care unit when compared to patients only admitted to the ward.

**Figure 2:**
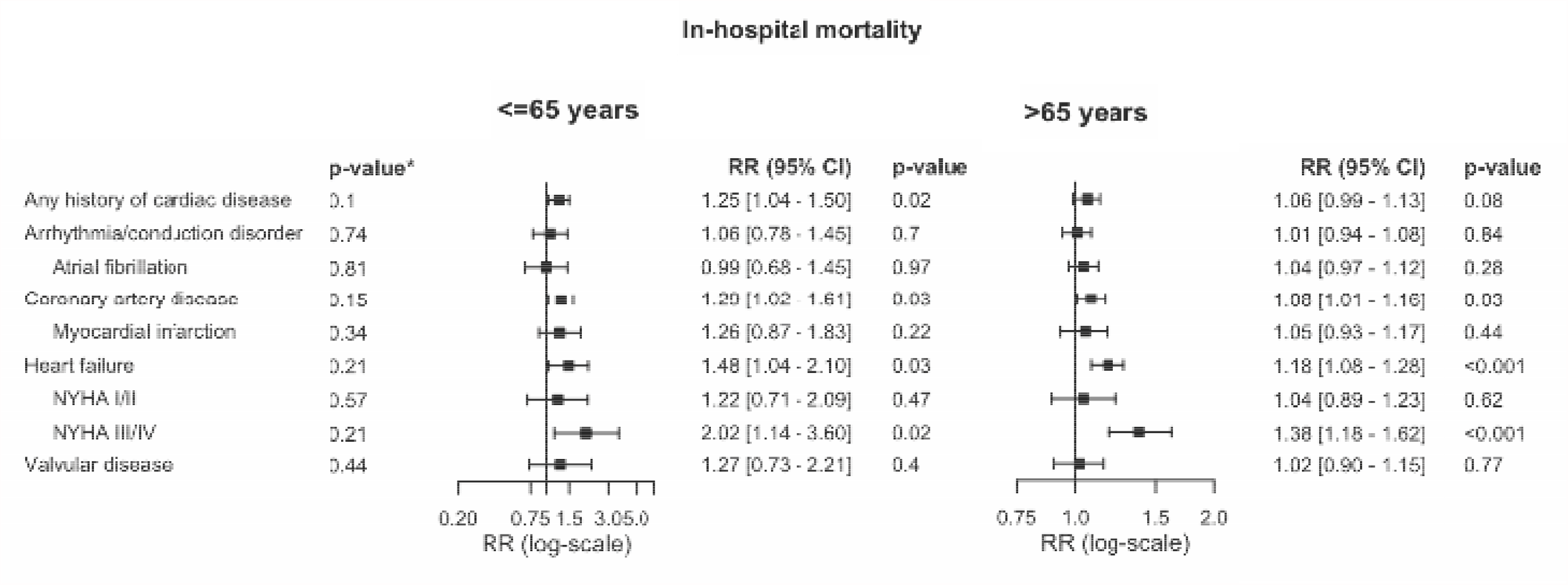
Multivariable adjusted associations between any history of cardiac disease, heart disease subtypes and in-hospital mortality in patients aged ≤65 (top) and >65 years (bottom). **\***p-value interaction heart disease subtype and age. Patients with prior myocardial infarction are included in the coronary artery disease group.

## Discussion

A large proportion of patients developing severe COVID-19 have underlying CVD. The aim of this study was to describe and compare the disease course and outcomes in hospitalized COVID-19 patients with and without pre-existing cardiac disease. The most important findings of this work are (i) the symptoms of COVID-19 at presentation are age dependent and do not differ in individuals with and without prior cardiac disease (ii) serious incident cardiac complications are diagnosed infrequently during hospitalization and are seen more often in patients with known cardiac disease at baseline (iii) the association between prior heart disease and in-hospital mortality varies across heart disease subtypes where (iv) heart failure associates most strongly with in-hospital mortality and (v) there was no association with ischemic heart disease.

In line with others^28^, we found that COVID-19 symptoms at admission vary mainly with age with no evidence that prior cardiac disease influences this relationship or the symptoms. Typical symptoms of COVID- 19 including fever, cough and shortness of breath, were reported less often in the elderly. Overall complaints mimicking cardiac disease, such as chest pain, palpitations and orthopnea were reported by only a minority of patients (<10%). In this regard, it is interesting that cardiac-like symptoms such as chest pain and palpitations have been reported by 17-44% and 20-32% of patients in the 2-3 months after the active infection as part of the so-called “long COVID” syndrome.^29-32^ Whether these complaints are related to cardiovascular involvement in the convalescent phase of the disease or should predominantly be viewed as an epiphenomenon remains unclear.

According to our study, serious cardiac complications are rarely diagnosed during hospitalization with COVID-19, with a prevalence <2% of patients. To aid interpretation of these numbers it is of interest to relate them to the occurrence of cardiac complications in patients hospitalized for other (viral) infectious diseases. A recent large French retrospective cohort study found that the prevalence of MI and atrial fibrillation was lower in patients with COVID-19 than seasonal influenza with a prevalence of 0.6% vs. 1.1% and 12.4% vs. 15.8% respectively.^7^ Unfortunately, this study did not report the prevalence of any other cardiac complications, including heart failure and myocarditis. As cardiomyocytes express ACE2^33,34^, the docking receptor of SARS-CoV-2, it has been speculated that SARS-CoV-2 may infect cardiomyocytes, replicate in cardiac tissue and thereby induce direct myocardial damage. These concerns have also been triggered by the finding that raised levels of troponin above test-specific upper limits are found in up to one third of patients at hospital admission.^35,36^ Histopathological studies of myocardial tissue in the setting of COVID-19 have been scarce up to this point. A literature review evaluating findings of 22 studies across 277 post-mortem examinations, found evidence for myocarditis in <2%.^37^ This finding contrasts the results of a number of cardiac magnetic resonance (CMR) studies evaluating tissue characteristics and function 2 – 3 months after an established SARS-CoV-2 infection.^32,38-40^ In the largest study by Kotecha et al. among 148 patients that had a troponin elevation during hospitalization, non-ischemic myocarditis-like late gadolinium enhancement was found in 26% of patients with one-third showing signs of active myocarditis.^39^ These findings were not associated with left ventricular dysfunction. In CMR studies conducted predominantly among clinically recovered mildly symptomatic or asymptomatic cases, up to 60% were described to have raised native T2 times, and which the authors suggested might be due to ongoing myocardial inflammation.^32,40^ Whether these CMR findings are unique to patients that have been infected with SARS-CoV-2, or also are seen in other (viral) infectious diseases has been poorly investigated. Evidence of a clear causal relationship between SARS-CoV-2 and myocarditis thereby remains elusive. It can be speculated that most patients may have troponin elevations secondary to profound hypoxia and a supply/demand imbalance rather than direct damage due to viral invasion in cardiac tissue.^36^ However, the discrepancy in the limited number of patients diagnosed with cardiac complications during hospitalization and the significant proportion of patients with abnormal findings in imaging studies is a concern and warrants further investigation. Cardiac complications may have been missed, due to the overlapping symptomatology with COVID-19 as well as limited access or performed diagnostic tests.

Contrary to cardiac complications, which in our study were rarely diagnosed during hospitalization, venous thrombosis and thromboembolism are common features of COVID-19, with a prevalence 3-4 times higher when compared to seasonal influenza.^7^ Thromboembolic events are especially common in patients admitted to the ICU, with pulmonary embolism being diagnosed more than 5 times as often (10.5% vs. 1.4%) in our cohort compared to patients treated on the ward only. The prevalence of pulmonary embolism in the ICU population in our study is lower than in studies based on data originating from patients hospitalized in the first months of the pandemic, that reported a prevalence of up to 20.6%.^41,42^ This discrepancy most likely reflects the implementation of enhanced antithrombotic prophylactic strategies in patients admitted with COVID-19 during 2020 and 2021.^43,44^ Interestingly, we observed that thromboembolic complications were less common among patients known with cardiac disease in our cohort. This observation is possibly related to pre-admission use of anticoagulants for the treatment of pre-existing cardiac conditions. This will be explored in ongoing analyses.

Among different heart disease subtypes, heart failure and especially severe heart failure (NYHA III/IV) was most strongly associated with in-hospital mortality in this study across the spectrum of different heart disease subtypes. Others have also identified patients with heart failure as one of the groups at particular risk.^45,46^ Among 6,439 hospitalized patients, in-hospital mortality was significantly higher among patients with a history of heart failure (aOR 1.88 [95% CI 1.27 – 2.78]).^45^ Similar findings were reported by the Tomasoni et al. (n=692) with a crude HR of 2.43 [95% CI 1.69 – 3.50] for heart failure remaining significant after adjustment for age, sex, various comorbidities and vitals and laboratory values at admission (adjusted HR 2.25 [95% CI 1.26 – 4.02].^46^ Whether the absolute risk of being hospitalized in the presence of heart failure is also increased was recently investigated in a population-based study, which found an aOR of 4.43 (95% CI 2.59 – 8.04; p- value <0.001).^47^ Besides age and male sex, heart failure had the strongest association with in-hospital mortality among various different comorbidities in this study.

Importantly, besides heart failure, none of the other types of heart disease were associated with in-hospital mortality after adjustment for age, sex, BMI, diabetes, hypertension, CKD and COPD. This heterogeneity was also evident in the population based study by Petrilli et al. in which patients with CAD did not seem to be at increased risk of hospitalization due to COVID-19 (aOR was 1.08; 95% CI 0.81 – 1.44; p- value 0.60).^47^ It could therefore be questioned whether all patients with heart disease should be defined as a group at risk, certainly when viewed in context of other demographic factors such as age and sex, as these appear to contribute to COVID-19 outcome to a much larger extent than pre-existing cardiac disease.^12^ This finding is of relevance for clinicians in countries with low vaccination rates and limited critical care capacity, that sometimes are forced to strict prioritization of the initiation and continuation of critical care treatment during this pandemic. Based on the results of this study, a history of cardiac disease, besides severe heart failure, should on itself presumably not be a reason to refrain from critical care treatment.

As heart failure and atrial fibrillation are primarily diseases of the elderly, with a steep increase in the prevalence as well as severity beyond the age of 75 years^48-50^, a majority of patients with the heart disease subtypes associated with the highest risk as identified in this study are among the first to have been vaccinated according to current vaccine rollout strategies across Europe.^27^ Adults with comorbidities are identified as an additional priority group but current advice lacks detail on which comorbidities should be considered high-risk. Therefore, we also determined the strength per heart disease subtype and in-hospital mortality in patients <65 years. Due to the lower comorbidity burden in this group, we hypothesized that the associations for the different types of heart disease would be higher in this patient population. However, due to the limited prevalence of heart disease in those aged <65 years of age, we lacked sufficient power to draw any strong conclusions based on this analysis.

### Limitations

Our study determines associations on a population level (e.g., all patients with heart failure) rather than individual risks and is limited to patients with COVID-19 that were hospitalized. In some countries that provided data, hospitalization and potentially life-sustaining treatments such as mechanical ventilation, might have been withheld in those with high frailty, including those with severe heart failure, which may have led to an overestimation of the found associations. Conversely, in younger patients treated more intensively, the associations of pre-existing cardiac disease with in-hospital mortality may be underestimated. This heterogeneity may have an impact the found associations. Furthermore, we could not reliably investigate associations between heart disease subtypes and HDU/ICU admission since we observed that patients admitted to a critical care unit were overall younger with fewer comorbidities. Since age in particular is well-known to be associated with a more severe COVID-19 disease course, this difference in baseline characteristics in those admitted to HDU/ICU is suggestive of an underlying selection of patients admitted for critical care. The mechanisms behind this selection are likely complex, influenced by amongst others the variability in critical care bed numbers across participating countries and available staff that may have led to demand for life-saving resources (nearly) exceeding supply, patient preference as well as cultural differences in clinical decision making. A further study limitation is that we only examined the impact of pre-existing cardiac disease on in-hospital mortality, which excludes the many deaths that have occurred in community settings and nursing homes. Moreover, there was no central adjudication of events in either of the two registries. Finally, echocardiographic data, providing more in-depth insight into heart failure etiology as well as the degree of systolic and diastolic dysfunction prior to hospitalization are lacking.

### Future perspectives

With the approval of several effective vaccines for COVID-19 there is hope of starting the containment of COVID-19 during 2021. During the first year of the pandemic, the scientific community has improved the understanding of this new disease considerably, including its effects on the cardiovascular system. However, many aspects are still unknown. Evidence for a strong causal relationship between SARS-CoV-2 and myocarditis is still lacking. The discrepancy between the low prevalence of clinically diagnosed cardiac complications among the most ill requiring hospitalization and the large proportion of only mildly or even asymptomatic patients with abnormal findings on CMR after the acute phase of the disease has ceased warrant further investigation to understand their specificity and significance. In addition, studies on the long-term incidence of major adverse cardiac events (MACE) are required. Future studies should also evaluate the added value of different pre-existing cardiac comorbidities such as severe heart failure in prognostic models.

### Conclusion

In this large retrospective cohort study across sixteen countries, more than one in three patients hospitalized with COVID-19 had underlying chronic cardiac disease. Patients with a history of heart disease were older, more frequently male and had a higher burden of other comorbid conditions at baseline. Inherently, patients with a history of heart disease have a poorer outcome once infected with SARS-CoV-2. However, after multivariable adjustment we did not find a significant association between chronic heart disease and in-hospital mortality. When evaluating the associations between specific heart disease subtypes and in-hospital mortality, considerable heterogeneity was detected. Of all patients with heart disease, those with heart failure are at greatest risk of death when hospitalized with COVID-19. None of the other heart disease subtypes investigated was significantly associated with in-hospital mortality. Furthermore, besides pulmonary embolism, serious cardiovascular complications are rarely diagnosed during hospital admission.

## Data Availability

The data underlying this article cannot be shared publicly for the privacy of individuals that participated in the study. The data will be shared on reasonable request to the corresponding author.

## Acknowledgements

We want to express our gratitude and appreciation to all participating sites and researchers part of the CAPACITY-COVID collaborative consortium and LEOSS study and all research professionals that have contributed to the data collection. CAPACITY-COVID gratefully acknowledges the following organizations for their assistance in the development of the registry and/or coordination regarding the data registration in the collaborating centers: partners of the Dutch CardioVascular Alliance (DCVA), the Dutch Association of Medical Specialists (FMS) and the British Heart Foundation Centres of Research Excellence. In addition, the consortium is thankful for the endorsement of the CAPACITY-COVID initiative by the European Society of Cardiology (ESC), the European Heart Network (EHN) and the Society for Cardiovascular Magnetic Resonance (SCMR). Furthermore, the consortium appreciates the endorsmenet of CAPACITY-COVD as a flagship research project within the National Institute for Health Research (NIHR)/British Heart Foundation (BHF) Partnership framework for COVID-19 research. Part of this work is supported by the BigData@Heart Consortium, funded by the Innovative Medicines Initiative-2 joint undertaking under grant agreement no 116074. This joint undertaking receives support from the EU’s Horizon 2020 research and innovation programme and EFPIA. Furthermore we want to thank the LEOSS study infrastructure group: Jörg Janne Vehreschild (Goethe University Frankfurt, University Hospital of Cologne), Lisa Pilgram (Goethe University Frankfurt), Carolin E. M. Jakob (University Hospital of Cologne), Melanie Stecher (University Hospital of Cologne), Max Schons (University Hospital of Cologne), Susana M. Nunes de Miranda (University Hospital of Cologne), Annika Claßen (University Hospital of Cologne), Sandra Fuhrmann (University Hospital of Cologne), Bernd Franke (University Hospital of Cologne), Nick Schulze (University Hospital of Cologne), Fabian Praßer (Charité, Universitätsmedizin Berlin) and Martin Lablans (University Medical Center Mannheim).

