## Supplemental Appendix for "Clinical presentation, disease course and outcome of COVID-19 in hospitalized patients with and without pre-existing cardiac disease – a cohort study across eighteen countries"

| Table S1: Contributing hospitals | |
|---|---|
| CAPACITY-COVID | |
| **Country** | **Site name** |
| Belgium | Antwerp University Hospital |
| | AZ Maria Middelares |
| | CHU UCL Namur – Site Godinne |
| | Jessa Hospital |
| | University Hospital Brussels |
| Egypt | One Day Surgery Hospital |
| France | SSR Val Rosay |
| Iran | Tehran Heart Center |
| Israel | EMMS Hospital |
| Italy | San Luigi Gonzaga University Hospital |
| Netherlands | Admiraal de Ruyter Hospital |
| | Albert Schweitzer Hospital |
| | Amphia Hospital |
| | Amstelland Hospital |
| | Amsterdam University Medical Center |
| | Antonius Hospital |
| | Beatrix Hospital |
| | Bernhoven Hospital |
| | Bravis Hospital |
| | Catharina Hospital |
| | Deventer Hospital |
| | Diakonessenhuis |
| | Dijklander Hospital, location Hoorn |
| | Elizabeth-TweeSteden Hospital |
| | Erasmus University Medical Center |
| | Franciscus Gasthuis |
| | Franciscus Vlietland |
| | Gelre Hospitals, location Apeldoorn |
| | Gelre Hospitals, location Zutphen |
| | Groene Hart Hospital |
| | Haaglanden Medical Center |
| | Hospital Group Twente |
| | Ikazia Hospital |
| | Isala |
| | Jeroen Bosch Hospital |
| | LangeLand Hospital |
| | Leiden University Medical Center |
| | Maasstad Hospital |
| | Maastricht University Medical Center |
| | Martini Hospital |
| | Meander Medical Center |
| | Medical Center Leeuwarden |
| | Medisch Spectrum Twente |
| | Rijnstate Hospital |
| | Rode Kruis Hospital |
| | Saxenburgh Medical Center |
| | Slingeland Hospital |
| | Spaarne Gasthuis |
| | St. Antonius Hospital |
| | St. Jansdal |
| | Treant Zorggroep |
| | University Medical Center Utrecht |
| | van Weel-Bethesda Hospital |
| | Zaans Medical Center |
| | Zuyderland Medical Center |
| Portugal | Hospital do Espirito Santo |
| | Hospital Prof. Doutor Fernando Fonesca |
| Russia | I.M. Sechenov First Moscow State Medical University |

| | |
|---|---|
| Saudi Arabia | King Fahd Hospital of the University |
| Spain | INCLIVA Research Institute, University of Valencia |
| | University Hospital Complex of Granada |
| Switzerland | University Hospital of Geneva |
| United Kingdom | Barts Health NHS Trust |
| | Leeds Teaching Hospitals NHS Trust |
| | Northumbria Healthcare NHS Foundation Trust |
| | Royal Brompton and Harefield NHS Foundation Trust |
| | Royal Devon and Exeter NHS Foundation Trust |
| | Royal Free London NHS Foundation Trust |
| | Salford Royal NHS Foundation Trust |
| | Southern Health and Social Care Trust |
| | The Newcastle upon Tyne Hospitals NHS Foundation Trust |
| | University College London Hospitals NHS Foundation Trust |
| | University Hospitals Bristol NHS Foundation Trust |
| | University Hospitals of Leicester NHS Trust |
| **LEOSS** | |
| **Country** | **Site name** |
| Austria | Medical University Graz |
| Belgium | National MS Center Melsbroek |
| Bosnia and Herzegovina | Clinic for Infectious Diseases University Clinical Hospital Mostar |
| Germany | Agaplesion Diakonie Hospital Rotenburg |
| | ARCIM Institute at Filderklinik |
| | Bethesda Hospital Bergedorf |
| | Bundeswehr Hospital Koblenz |
| | Catholic Hospital Bochum (St. Josef Hospital) Ruhr University Bochum |
| | Clinic Munich |
| | Department of Nephrology and Internal Intensive Care Medicine, Charité Universitaetsmedizin Berlin |
| | Elbland Hospital Riesa |
| | Elisabeth Hospital Essen |
| | Evangelisches Hospital Herne |
| | Evangelisches Hospital Saarbruecken |
| | German Heart Center Munich |
| | Hegau-Bodensee Hospital Singen |
| | Helios Hospital Pirna |
| | Hospital Braunschweig |
| | Hospital Bremen-Center |
| | Hospital Dortmund gGmbH |
| | Hospital Ernst von Bergmann |
| | Hospital Fulda |
| | Hospital Ingolstadt |
| | Hospital Kreuznacher Diakonie Hunsrueck |
| | Hospital Leverkusen |
| | Hospital Maria Hilf GmbH Moenchengladbach |
| | Hospital Mutterhaus Borromaeerinnen Trier |
| | Hospital Nuremberg North |
| | Hospital of the Augustinian Cologne |
| | Hospital Oldenburg University Oldenburg |
| | Hospital Osnabrueck |
| | Hospital Passau |
| | Hospital Preetz |
| | Hospital Saar Sulzbach |
| | Hospital Sankt Georg Leipzig |
| | Hospital South-Eastern Bavaria Trostberg |
| | Hospital St. Josef Wiesbaden |
| | Hospital St. Joseph-Stift Dresden |
| | Hospital Stuttgart |
| | Hospital zum Heiligen Geist, Kempen |
| | Hospitals of Cologne gGmbH |

| | |
|---|---|
| | Johannes Wesling Hospital Minden Ruhr University Bochum |
| | Maltes Hospital St. Hildegardis Cologne |
| | Malteser Hospital St. Franziskus Flensburg |
| | Marien Hospital Herne Ruhr University Bochum |
| | Medical practice U. Kronawitter & C. Jung Oncology Traunstein |
| | Medical School Hannover |
| | Municipal Hospital Karlsruhe |
| | Nephrological Center Villingen-Schwenningen |
| | Oberlausitz Hospital |
| | Otto-von-Guericke-University Magdeburg |
| | Petrus Hospital Wuppertal |
| | Practice at Ebertplatz Cologne |
| | Practice Dr. Boebel Reutlingen |
| | Practice for general medicine Drs. Elisabeth Schroedter & Gabriele Mueller-Joerger |
| | Practice Gotenring Cologne |
| | Robert-Bosch-Hospital Stuttgart |
| | Robert-Koch-Institute |
| | Sankt Vincenz Hospital Menden |
| | Schwerpunktpraxis Onkologie und Haematologie |
| | SHG Clinics Voelklingen Lung Center Saar |
| | Sophien- and Hufeland Clinic Weimar |
| | Sophien Hospital GmbH Hannover |
| | Srh Wald-Hospital Gera |
| | St. Vincenz Hospital Datteln |
| | St. Vincenz Hospital Limburg/Lahn |
| | Technical University of Munich |
| | Thorax-Hospital Heidelberg |
| | Tropical Clinic Paul-Lechler Hospital Tuebingen |
| | University Heart Center Freiburg Bad Krozingen |
| | University Hospital Augsburg |
| | University Hospital Bonn |
| | University Hospital Cologne |
| | University Hospital Dresden |
| | University Hospital Duesseldorf |
| | University Hospital Erlangen |
| | University Hospital Essen |
| | University Hospital Frankfurt |
| | University Hospital Freiburg |
| | University Hospital Goettingen |
| | University Hospital Hamburg-Eppendorf |
| | University Hospital Heidelberg |
| | University Hospital Jena |
| | University Hospital Muenster |
| | University Hospital Munich/ LMU |
| | University Hospital of Giessen and Marburg |
| | University Hospital Regensburg |
| | University Hospital Rostock |
| | University Hospital RWTH Achen |
| | University Hospital Saarland |
| | University Hospital Schleswig-Holstein - Kiel |
| | University Hospital Schleswig-Holstein - Luebeck |
| | University Hospital Tuebingen |
| | University Hospital Ulm |
| | University Hospital Wuerzburg |
| | University Medicine of the Johannes Gutenberg University Mainz |
| | Zeisigwald Hospitals Bethanien Chemnitz |
| Italy | Sant' Andrea Hospital University of Rome |
| Latvia | P. Stradins University Hospital |
| Spain | Complejo Hospitalario de Navarra |
| | University Hospital Arnau de Vilanova Lleida |

| | |
|---|---|
| Switzerland | Department of Neurology, Inselspital, Bern University Hospital |
| | Kantonsspital St. Gallen |
| | Luzerner Kantonsspital Neurology |
| | Neurocenter of Southern Switzerland, EOC |
| | University Hospital Basel |
| Turkey | Hacettepe University |
| | Pamukkale University School of Medicine |
| | University of Health Sciences Istanbul |

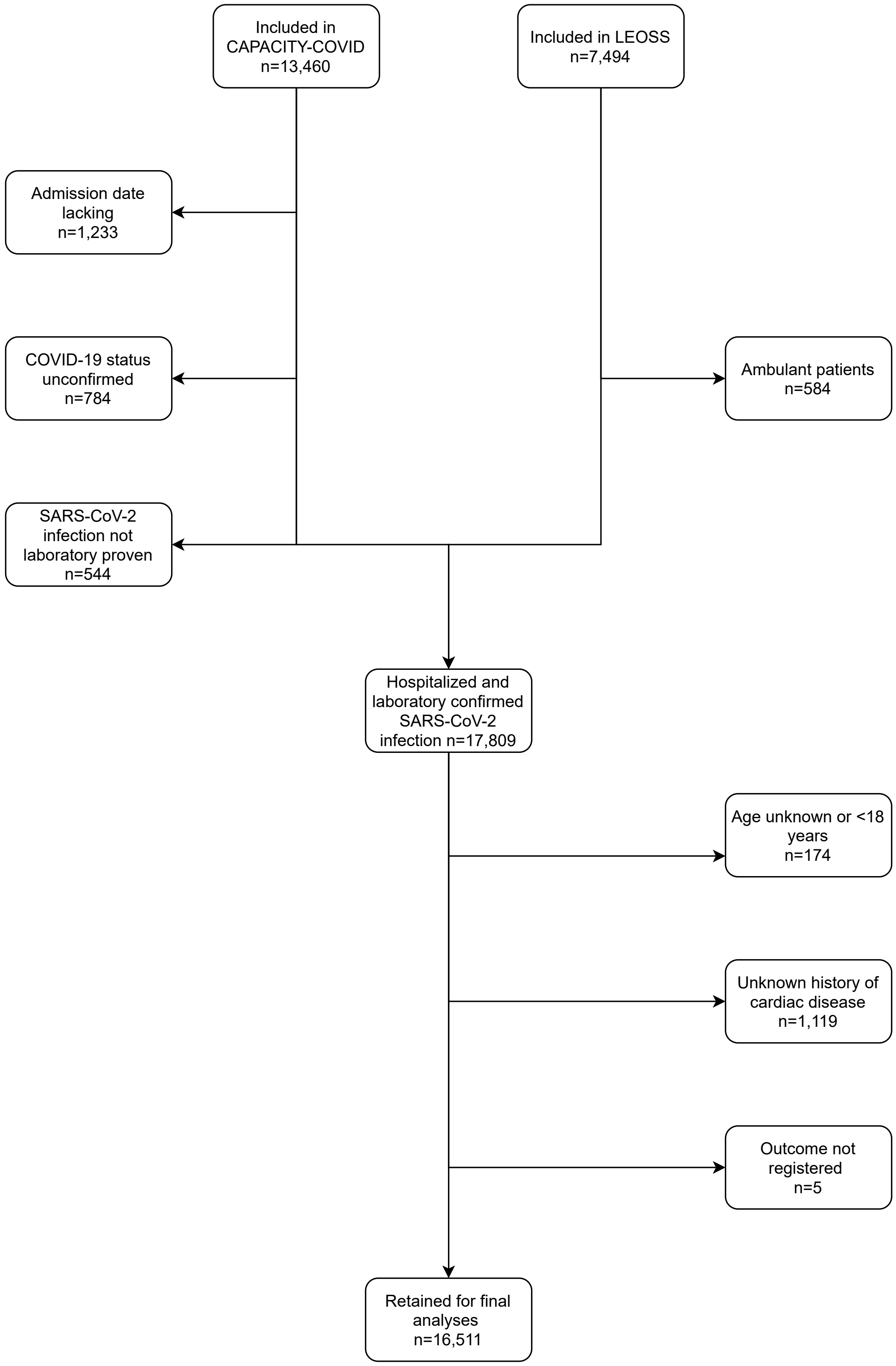

**Figure S1**: Selection of study population

| Table S2: Baseline characteristics stratified by data source and age | | | | | | |
|---|---|---|---|---|---|---|
| | Overall | CAPACITY-COVID | | LEOSS | | P-value |
| | | ≤65 years | >65 years | ≤65 years | >65 years | |
| Total | 16511 | 4303 | 5501 | 3347 | 3360 | |
| **Age, years (%)** | | | | | | <0.001 |
| Median, [IQR] | NA | 55 [46 – 60] | 77 [72 – 84] | NA | NA | |
| 18 – 25 | 239 (1.4) | 94 (2.2) | 0 (0.0) | 145 (4.3) | 0 (0.0) | |
| 26 – 35 | 704 (4.3) | 303 (7.0) | 0 (0.0) | 401 (12.0) | 0 (0.0) | |
| 36 – 45 | 1128 (6.8) | 581 (13.5) | 0 (0.0) | 547 (16.3) | 0 (0.0) | |
| 46 – 55 | 2299 (13.9) | 1309 (30.4) | 0 (0.0) | 990 (29.6) | 0 (0.0) | |
| 56 – 65 | 3280 (19.9) | 2016 (46.9) | 0 (0.0) | 1264 (37.8) | 0 (0.0) | |
| 66 – 75 | 3485 (21.1) | 0 (0.0) | 2290 (41.6) | 0 (0.0) | 1195 (35.6) | |
| 76 – 85 | 3720 (22.5) | 0 (0.0) | 2178 (39.6) | 0 (0.0) | 1542 (45.9) | |
| >85 | 1656 (10.0) | 0 (0.0) | 1033 (18.8) | 0 (0.0) | 623 (18.5) | |
| **Sex, female (%)** | 6627 (40.2) | 1559 (36.3) | 2222 (40.5) | 1311 (39.2) | 1535 (45.7) | <0.001 |
| **Ethnicity** | | | | | | <0.001 |
| Arab | 496 (3.5) | 345 (9.0) | 151 (3.0) | 0 (0.0) | 0 (0.0) | |
| Asian | 898 (6.3) | 483 (12.6) | 231 (4.6) | 143 (5.5) | 41 (1.4) | |
| Black | 384 (2.7) | 215 (5.6) | 85 (1.7) | 78 (3.0) | 6 (0.2) | |
| Latin-American | 21 (0.1) | 14 (0.4) | 7 (0.1) | 0 (0.0) | 0 (0.0) | |
| White | 12120 (84.5) | 2481 (64.8) | 4464 (88.2) | 2358 (91.4) | 2817 (98.4) | |
| Other | 416 (2.9) | 293 (7.6) | 123 (2.4) | 0 (0.0) | 0 (0.0) | |
| **BMI (kg/m$^2$)(%)** | | | | | | <0.001 |
| Mean, (+/- SD) | NA | 29.5 (6.3) | 27.2 (5.6) | NA | NA | |
| Underweight (<18.5) | 239 (2.3) | 22 (0.8) | 118 (3.3) | 36 (1.7) | 63 (3.2) | |
| Normal weight (18.5 – 24.9) | 3091 (29.7) | 601 (22.3) | 1205 (33.3) | 580 (27.6) | 705 (35.3) | |
| Overweight (25.0 – 29.9) | 3854 (37.0) | 1027 (38.1) | 1365 (37.7) | 753 (35.8) | 709 (35.5) | |
| Obese (30.0 – 34.9) | 2069 (19.9) | 619 (23.0) | 643 (17.8) | 450 (21.4) | 357 (17.9) | |
| Morbidly obese (> 34.9) | 1157 (11.1) | 424 (15.7) | 288 (8.0) | 283 (13.5) | 162 (8.1) | |
| **Cardiovascular risk factors (%)** | | | | | | |
| Diabetes | 4031 (24.9) | 886 (21.0) | 1649 (30.4) | 494 (15.0) | 1002 (30.5) | <0.001 |
| Hypertension | 7975 (49.5) | 1310 (31.2) | 3232 (60.3) | 1089 (33.2) | 2344 (71.5) | <0.001 |
| Peripheral arterial disease | 654 (4.8) | 49 (1.5) | 284 (7.1) | 53 (1.6) | 268 (8.3) | <0.001 |
| **Cardiac disease (%)** | | | | | | |
| Any history of cardiac disease | 5198 (31.5) | 654 (15.2) | 2647 (48.1) | 345 (10.3) | 1552 (46.2) | <0.001 |
| Arrhythmia/conduction disorder | 2503 (15.3) | 174 (4.0) | 1277 (23.2) | 113 (3.4) | 939 (28.4) | <0.001 |
| Atrial fibrillation/flutter | 2004 (12.3) | 96 (2.2) | 953 (17.3) | 100 (3.0) | 855 (26.1) | <0.001 |
| Coronary artery disease | 2420 (14.8) | 284 (6.6) | 1090 (19.8) | 225 (6.8) | 821 (25.1) | <0.001 |
| Myocardial infarction | 769 (4.8) | 73 (1.7) | 298 (5.6) | 102 (3.1) | 296 (9.1) | <0.001 |
| Heart failure | 1314 (8.1) | 86 (2.0) | 630 (11.5) | 84 (2.6) | 514 (15.9) | <0.001 |
| NYHA I/II | 324 (2.0) | 26 (0.6) | 157 (2.9) | 24 (0.7) | 117 (3.5) | <0.001 |
| NYHA III/IV | 246 (1.5) | 4 (0.1) | 75 (1.4) | 20 (0.6) | 147 (4.4) | <0.001 |
| Valvular heart disease | 579 (3.6) | 53 (1.2) | 368 (6.7) | 18 (0.6) | 140 (4.3) | <0.001 |
| **Comorbidities (%)** | | | | | | |
| Chronic kidney disease | 2196 (13.5) | 189 (4.4) | 991 (18.2) | 223 (6.8) | 793 (24.2) | <0.001 |
| COPD | 1563 (9.6) | 245 (5.8) | 887 (16.4) | 98 (3.0) | 333 (10.2) | <0.001 |
| **Use cardiovascular drugs (%)** | | | | | | |
| ACE-inhibitors | 3021 (18.8) | 491 (11.4) | 1144 (20.8) | 410 (13.1) | 976 (31.2) | <0.001 |
| Aldosterone antagonist | 563 (3.5) | 53 (1.2) | 219 (4.0) | 63 (1.9) | 228 (7.1) | <0.001 |
| Anti-platelet | 3044 (18.7) | 417 (9.7) | 1419 (25.8) | 274 (8.4) | 934 (29.1) | <0.001 |
| Angiotensin receptor blocker | 2159 (13.5) | 316 (7.4) | 745 (13.6) | 343 (11.0) | 755 (24.4) | <0.001 |
| Calcium-blocker | 2769 (17.0) | 503 (11.7) | 1108 (20.2) | 365 (11.2) | 793 (24.7) | <0.001 |
| Diuretic | 3338 (20.5) | 395 (9.2) | 1515 (27.6) | 291 (9.0) | 1137 (35.4) | <0.001 |
| Insulin | 1204 (7.4) | 278 (6.5) | 431 (7.8) | 146 (4.4) | 349 (10.6) | <0.001 |
| Lipid-lowering | 4789 (30.5) | 911 (21.2) | 2459 (44.7) | 363 (12.3) | 1056 (35.6) | <0.001 |
| Oral anti-diabetic | 2139 (13.0) | 523 (12.2) | 973 (17.7) | 235 (7.0) | 408 (12.1) | <0.001 |

ACE = Angiotensin Converting Enzyme; BMI = Body Mass Index; COPD = Chronic Obstructive Pulmonary Disease; IQR = Interquartile Range; NYHA = New York Heart Association; SD = Standard Deviation

| Table S3: Detailed specification of pre-existing cardiac disease in CAPACITY COVID | | | | |
|---|---|---|---|---|
| | **Overall** | **≤65 years** | **>65 years** | **P-value** |
| Total | 9804 | 4303 | 5501 | |
| **Arrhythmia/conduction disorder** | | | | |
| Any | 1451 (14.8) | 174 (4.0) | 1277 (23.2) | <0.001 |
| Supraventricular tachycardia | 1179 (12.0) | 130 (3.0) | 1049 (19.1) | <0.001 |
| Paroxysmal atrial fibrillation | 442 (4.5) | 48 (1.1) | 394 (7.2) | <0.001 |
| Persistent atrial fibrillation | 183 (1.9) | 14 (0.3) | 169 (3.1) | <0.001 |
| Permanent atrial fibrillation | 378 (3.9) | 27 (0.6) | 351 (6.4) | <0.001 |
| Atrial flutter | 989 (10.1) | 89 (2.1) | 900 (16.4) | <0.001 |
| Atrial tachycardia | 87 (0.9) | 7 (0.2) | 80 (1.5) | <0.001 |
| AVNRT | 23 (0.2) | 10 (0.2) | 13 (0.2) | 1.00 |
| AVRT | 2 (0.0) | 1 (0.0) | 1 (0.0) | 1.00 |
| Unspecified | 91 (0.9) | 17 (0.4) | 74 (1.3) | <0.001 |
| Ventricular tachycardia (VT) | 66 (0.7) | 19 (0.4) | 47 (0.9) | 0.018 |
| Non-sustained VT | 24 (0.2) | 8 (0.2) | 16 (0.3) | 0.402 |
| Sustained VT | 19 (0.2) | 4 (0.1) | 15 (0.3) | 0.076 |
| Ventricular fibrillation | 26 (0.3) | 8 (0.2) | 18 (0.3) | 0.249 |
| Conduction disorder | 154 (1.6) | 13 (0.3) | 141 (2.6) | <0.001 |
| First-degree AV-block | 34 (0.3) | 3 (0.1) | 31 (0.6) | <0.001 |
| Second-degree AV-block | 19 (0.2) | 1 (0.0) | 18 (0.3) | 0.002 |
| Third-degree AV-block | 31 (0.3) | 0 (0.0) | 31 (0.6) | <0.001 |
| Left bundle branch block | 32 (0.3) | 6 (0.1) | 26 (0.5) | 0.007 |
| Right bundle branch block | 28 (0.3) | 0 (0.0) | 28 (0.5) | <0.001 |
| Long-QT | 7 (0.1) | 1 (0.0) | 6 (0.1) | 0.231 |
| Unspecified | 14 (0.1) | 3 (0.1) | 11 (0.2) | 0.154 |
| Sinus node dysfunction | 52 (0.5) | 5 (0.1) | 47 (0.9) | <0.001 |
| **Congenital heart disease** | 37 (0.4) | 18 (0.4) | 19 (0.3) | 0.676 |
| **Coronary artery disease** | 1374 (14.0) | 284 (6.6) | 1090 (19.8) | <0.001 |
| Stable angina | 385 (4.0) | 75 (1.8) | 310 (5.8) | <0.001 |
| Unstable angina | 197 (2.0) | 44 (1.0) | 153 (2.9) | <0.001 |
| NSTEMI | 371 (3.9) | 73 (1.7) | 298 (5.6) | <0.001 |
| STEMI | 361 (3.7) | 77 (1.8) | 284 (5.3) | <0.001 |
| **Intervention coronary artery disease** | | | | |
| PCI | 637 (6.6) | 142 (3.3) | 495 (9.1) | <0.001 |
| CABG | 366 (3.8) | 58 (1.4) | 308 (5.7) | <0.001 |
| **Heart failure** | 716 (7.3) | 86 (2.0) | 630 (11.5) | <0.001 |
| NYHA I/II | 183 (1.9) | 26 (0.6) | 157 (2.9) | <0.001 |
| NYHA III/IV | 79 (0.8) | 4 (0.1) | 75 (1.4) | <0.001 |
| **Cardiomyopathy** | | | | |
| Arrhythmogenic | 36 (0.4) | 4 (0.1) | 32 (0.6) | <0.001 |
| Dilated | 136 (1.4) | 12 (0.3) | 124 (2.3) | <0.001 |
| Hypertensive | 55 (0.6) | 8 (0.2) | 47 (0.9) | <0.001 |
| Hypertrophic | 21 (0.2) | 6 (0.1) | 15 (0.3) | 0.224 |
| Ischemic | 118 (1.2) | 20 (0.5) | 98 (1.8) | <0.001 |
| Myocarditis | 6 (0.1) | 3 (0.1) | 3 (0.1) | 1.000 |
| Non-compaction | 4 (0.0) | 0 (0.0) | 4 (0.1) | 0.203 |
| Restrictive | 4 (0.0) | 1 (0.0) | 3 (0.1) | 0.790 |
| Toxic | 4 (0.0) | 2 (0.0) | 2 (0.0) | 1.000 |
| Valvular | 58 (0.6) | 6 (0.1) | 52 (1.0) | <0.001 |
| Unspecified | 217 (2.2) | 15 (0.3) | 202 (3.7) | <0.001 |
| **Valvular heart disease** | 421 (4.3) | 53 (1.2) | 368 (6.7) | <0.001 |
| Aortic stenosis | 231 (2.4) | 25 (0.6) | 206 (3.7) | <0.001 |
| Aortic regurgitation | 66 (0.7) | 13 (0.3) | 53 (1.0) | <0.001 |
| Mitral stenosis | 26 (0.3) | 3 (0.1) | 23 (0.4) | 0.002 |
| Mitral regurgitation | 130 (1.3) | 16 (0.4) | 114 (2.1) | <0.001 |
| Pulmonary regurgitation | 3 (0.0) | 1 (0.0) | 2 (0.0) | 1.000 |
| Tricuspid regurgitation | 51 (0.5) | 3 (0.1) | 48 (0.9) | <0.001 |
| **Interventions valvular heart disease** | | | | |
| Intervention performed | 174 (1.8) | 26 (0.6) | 148 (2.7) | <0.001 |
| Intervention planned | 11 (0.1) | 3 (0.1) | 8 (0.1) | 0.420 |

AVNRT = AV-nodal Reentrant Tachycardia; AVRT = Atrioventricular Reentrant Tachycardia; AV = Atrioventricular; CABG = Coronary Artery Bypass Surgery; NSTEMI = Non-ST-Elevation Myocardial Infarction; NYHA = New York Heart Association; PCI = Percutaneous Coronary Intervention; STEMI = ST-Elevation Myocardial Infarction; VT = Ventricular Tachycardia

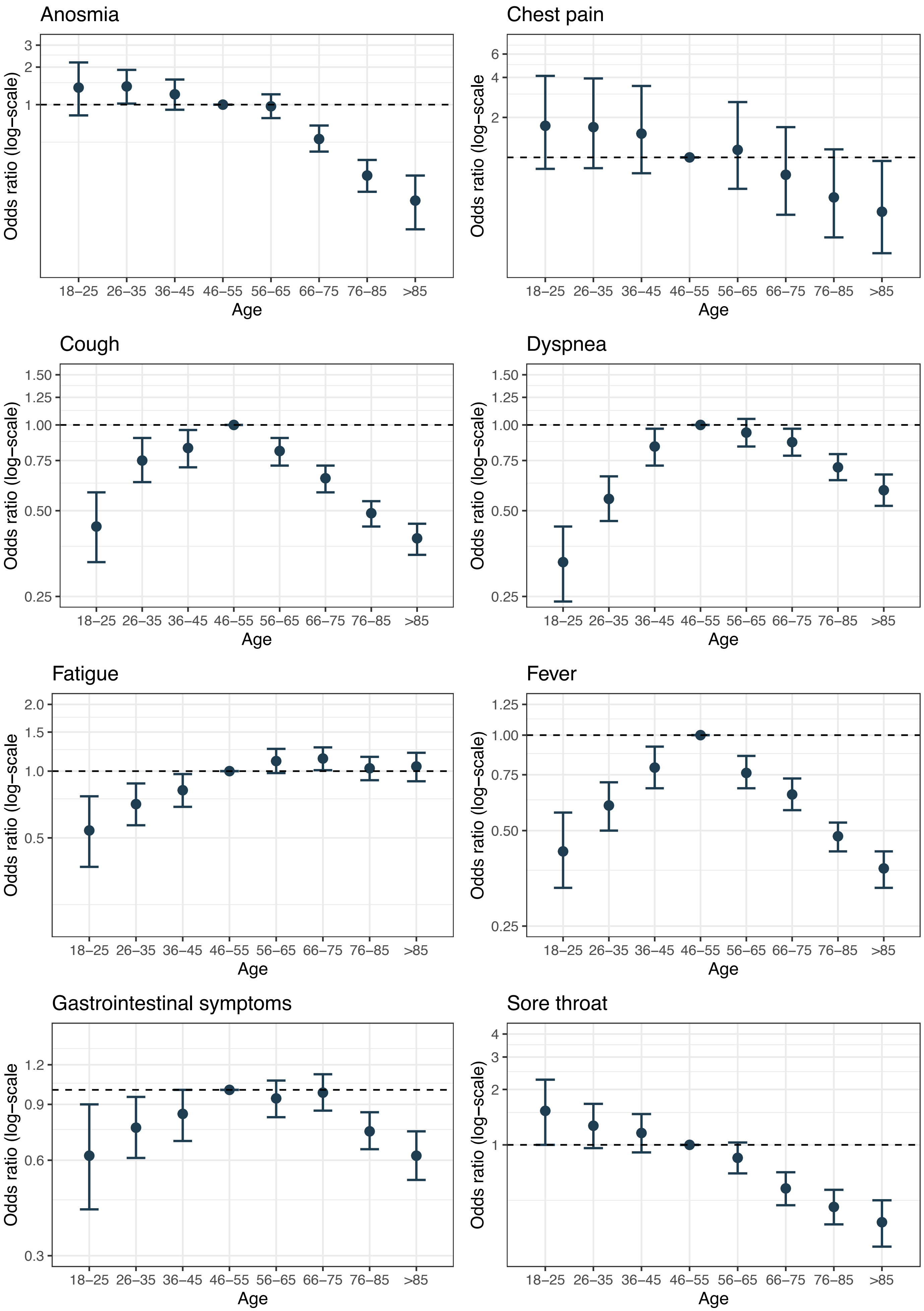

**Figure S2**: Complaints at presentation stratified by age.

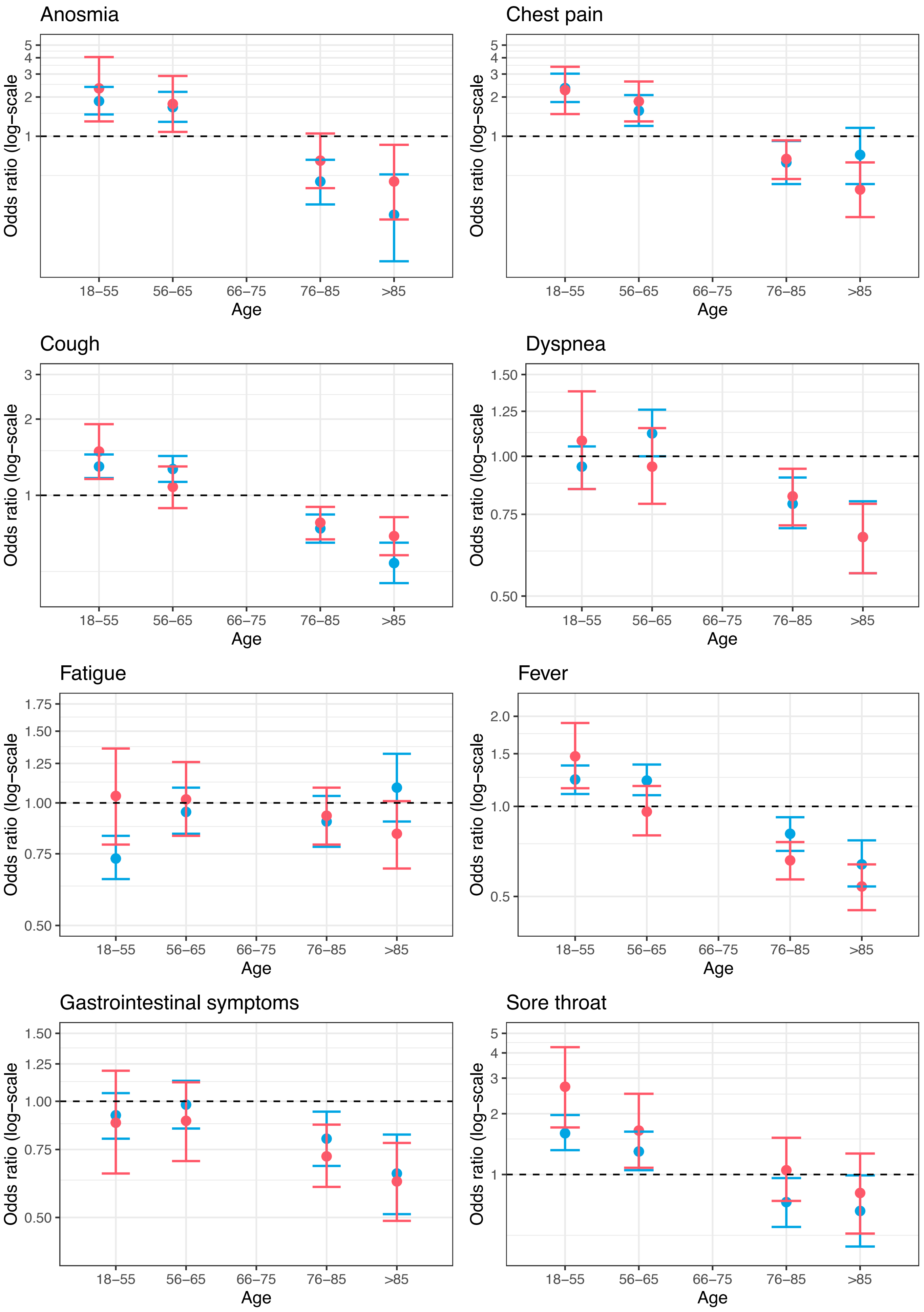

**Figure S3:** Complaints at presentation stratified by age and prior history of cardiac disease. Red = patients with pre-existing cardiac disease, blue = patients without pre-existing cardiac disease.

| Table S4: Complaints, vitals and laboratory values at admission stratified by data source and age | | | | | | |
|---|---|---|---|---|---|---|
| | **Overall** | **CAPACITY-COVID** | | **LEOSS** | | **P-value** |
| | | **≤65 years** | **>65 years** | **≤65 years** | **>65 years** | |
| Total | 16511 | 4303 | 5501 | 3347 | 3360 | |
| **Admission** | | | | | | |
| Onset of symptoms to admission, days, median, [IQR] | NA | 7 [3 – 10] | 5 [1 – 8] | NA | NA | NA |
| **Complaints at admission (%)** | | | | | | |
| Anosmia | 723 (4.6) | 297 (7.4) | 153 (2.9) | 217 (6.7) | 56 (1.7) | <0.001 |
| Chest pain | NA | 535 (13.3) | 318 (6.1) | NA | NA | NA |
| Cough | 8158 (51.8) | 2713 (67.5) | 2867 (55.1) | 1510 (46.5) | 1068 (32.7) | <0.001 |
| Dyspnea | 7798 (49.8) | 2694 (67.0) | 3027 (58.2) | 1056 (33.0) | 1021 (31.7) | <0.001 |
| Fatigue | 4214 (26.8) | 1399 (34.8) | 1790 (34.4) | 471 (14.5) | 554 (16.9) | <0.001 |
| Fever | 8638 (54.9) | 2914 (72.5) | 3104 (59.6) | 1491 (45.9) | 1129 (34.5) | <0.001 |
| Gastrointestinal symptoms | 3030 (19.2) | 1120 (27.9) | 1269 (24.4) | 352 (10.8) | 289 (8.8) | <0.001 |
| (Pre) syncope | NA | 108 (2.7) | 205 (3.9) | NA | NA | NA |
| Orthopnea | NA | 68 (1.7) | 66 (1.3) | NA | NA | NA |
| Palpitations | 162 (1.0) | 42 (1.0) | 62 (1.2) | 24 (0.7) | 34 (1.0) | 0.258 |
| Peripheral edema | NA | 13 (0.3) | 42 (0.8) | NA | NA | NA |
| Sore throat | 1108 (7.0) | 421 (10.5) | 317 (6.1) | 270 (8.3) | 100 (3.1) | <0.001 |
| **Vitals at admission (%)** | | | | | | |
| Temperature (°C) | | | | | | <0.001 |
| Mean, (SD) | NA | 37.7 (1.1) | 37.5 (1.1) | NA | NA | |
| < 35.1 | 72 (0.6) | 13 (0.3) | 42 (0.9) | 4 (0.3) | 13 (0.8) | |
| 35.1 – 37.2 | 4898 (41.7) | 1383 (36.3) | 2049 (43.4) | 665 (42.5) | 801 (48.9) | |
| 37.3 – 37.9 | 2546 (21.7) | 877 (23.0) | 1029 (21.8) | 313 (20.0) | 327 (20.0) | |
| 38.0 – 38.9 | 2920 (24.9) | 1031 (27.0) | 1141 (24.2) | 390 (24.9) | 358 (21.9) | |
| 39.0 – 39.9 | 1139 (9.7) | 451 (11.8) | 402 (8.5) | 164 (10.5) | 122 (7.4) | |
| > 39.9 | 158 (1.3) | 59 (1.5) | 54 (1.1) | 28 (1.8) | 17 (1.0) | |
| Respiratory rate (breaths/min) | | | | | | <0.001 |
| Median, [IQR] | NA | 21 [18 – 26] | 20 [18 – 25] | NA | NA | |
| < 16 | 1168 (10.8) | 267 (7.2) | 391 (8.5) | 246 (20.4) | 264 (20.0) | |
| 16 – 21 | 4914 (45.4) | 1646 (44.5) | 2059 (44.9) | 596 (49.5) | 613 (46.4) | |
| 22 – 29 | 3219 (29.8) | 1179 (31.9) | 1476 (32.2) | 249 (20.7) | 315 (23.9) | |
| > 29 | 1512 (14.0) | 609 (16.5) | 661 (14.4) | 114 (9.5) | 128 (9.7) | |
| Heart rate (beats/min) | | | | | | <0.001 |
| Mean, (SD) | NA | 93 (18) | 88 (20) | NA | NA | |
| <45 | 51 (0.4) | 10 (0.3) | 27 (0.6) | 7 (0.4) | 7 (0.4) | |
| 45 – 59 | 293 (2.5) | 48 (1.3) | 168 (3.5) | 30 (1.9) | 47 (2.9) | |
| 60 – 89 | 6037 (51.2) | 1601 (41.7) | 2548 (53.7) | 858 (55.1) | 1030 (62.5) | |
| 90 – 119 | 4627 (39.3) | 1881 (49.0) | 1671 (35.2) | 593 (38.1) | 482 (29.2) | |
| >119 | 775 (6.6) | 297 (7.7) | 327 (6.9) | 68 (4.4) | 83 (5.0) | |
| Systolic blood pressure (mmHg) | | | | | | <0.001 |
| Mean, (SD) | NA | 131 (21) | 134 (24) | NA | NA | |
| <80 | 79 (0.7) | 14 (0.4) | 37 (0.8) | 8 (0.5) | 20 (1.2) | |
| 80 – 99 | 566 (4.8) | 166 (4.3) | 259 (5.5) | 59 (3.8) | 82 (4.9) | |
| 100 – 119 | 2606 (22.1) | 920 (24.1) | 1028 (21.7) | 357 (22.9) | 301 (18.2) | |
| 120 – 139 | 4291 (36.5) | 1564 (40.9) | 1538 (32.5) | 653 (41.9) | 536 (32.3) | |
| 140 – 179 | 3827 (32.5) | 1077 (28.2) | 1670 (35.3) | 450 (28.9) | 630 (38.0) | |
| >179 | 403 (3.4) | 80 (2.1) | 205 (4.3) | 30 (1.9) | 88 (5.3) | |
| Diastolic blood pressure (mmHg) | | | | | | <0.001 |
| Mean, (SD) | NA | 79 (14) | 75 (15) | NA | NA | |
| < 40 | 44 (0.4) | 11 (0.3) | 17 (0.4) | 3 (0.2) | 13 (0.8) | |
| 40 – 59 | 1218 (10.4) | 278 (7.3) | 649 (13.7) | 84 (5.4) | 207 (12.6) | |
| 60 – 89 | 8452 (71.9) | 2773 (72.6) | 3352 (70.8) | 1174 (75.6) | 1153 (69.9) | |
| 90 – 109 | 1823 (15.5) | 696 (18.2) | 615 (13.0) | 266 (17.1) | 246 (14.9) | |
| > 109 | 216 (1.8) | 61 (1.6) | 100 (2.1) | 25 (1.6) | 30 (1.8) | |
| Oxygen saturation (%) | | | | | | <0.001 |
| Median, [IQR] | NA | 95 [93 – 97] | 95 [92 – 97] | NA | NA | |
| < 60 | 67 (0.6) | 21 (0.5) | 14 (0.3) | 12 (0.8) | 20 (1.2) | |
| 60 – 69 | 57 (0.5) | 19 (0.5) | 15 (0.3) | 7 (0.5) | 16 (1.0) | |

| | | | | | | |
|---|---|---|---|---|---|---|
| 70 – 79 | 206 (1.8) | 62 (1.6) | 67 (1.4) | 23 (1.5) | 54 (3.3) | |
| 80 – 89 | 1313 (11.2) | 347 (9.1) | 532 (11.2) | 141 (9.2) | 293 (17.9) | |
| 90 – 95 | 4768 (40.6) | 1492 (39.0) | 2050 (43.3) | 540 (35.1) | 686 (41.9) | |
| 96 – 100 | 5330 (45.4) | 1887 (49.3) | 2060 (43.5) | 816 (53.0) | 567 (34.7) | |
| **Laboratory values at admission (%)** | | | | | | |
| CRP (mg/L) | | | | | | <0.001 |
| Median, [IQR] | NA | 82 [36 – 157] | 84 [39 – 148] | NA | NA | |
| < 3 | 540 (4.7) | 81 (2.2) | 62 (1.4) | 251 (15.6) | 146 (8.6) | |
| 3 – 29 | 2664 (23.2) | 680 (18.7) | 803 (17.8) | 641 (39.9) | 540 (31.7) | |
| 30 – 69 | 2605 (22.7) | 844 (23.2) | 1078 (23.8) | 294 (18.3) | 389 (22.8) | |
| 70 – 119 | 2225 (19.4) | 738 (20.3) | 1005 (22.2) | 186 (11.6) | 296 (17.4) | |
| 120 – 179 | 1657 (14.4) | 562 (15.5) | 762 (16.9) | 148 (9.2) | 185 (10.9) | |
| 180 – 249 | 965 (8.4) | 375 (10.3) | 451 (10.0) | 50 (3.1) | 89 (5.2) | |
| > 249 | 812 (7.1) | 356 (9.8) | 360 (8.0) | 37 (2.3) | 59 (3.5) | |
| White blood cell count (x 10^9/L) | | | | | | <0.001 |
| Mean, (SD) | NA | 8.4 (10.1) | 8.5 (8.1) | NA | NA | |
| < 1.0 | 33 (0.3) | 12 (0.3) | 12 (0.3) | 7 (0.4) | 2 (0.1) | |
| 1.0 – 3.9 | 1327 (11.5) | 407 (11.1) | 430 (9.6) | 248 (15.1) | 242 (14.1) | |
| 4.0 – 7.9 | 6110 (53.0) | 1924 (52.3) | 2327 (51.9) | 961 (58.5) | 898 (52.2) | |
| 8.0 – 11.9 | 2773 (24.1) | 936 (25.4) | 1169 (26.1) | 308 (18.8) | 360 (20.9) | |
| 12.0 – 15.9 | 935 (8.1) | 306 (8.3) | 417 (9.3) | 79 (4.8) | 133 (7.7) | |
| 16.0 – 19.9 | 288 (2.5) | 93 (2.5) | 132 (2.9) | 17 (1.0) | 46 (2.7) | |
| > 20 | 60 (0.5) | 0 (0.0) | 0 (0.0) | 22 (1.3) | 38 (2.2) | |
| Lymphocyte count (x 10^9/L) | | | | | | <0.001 |
| Mean, (SD) | NA | 1.13 (0.83) | 1.01 (0.96) | NA | NA | |
| < 0.1 | 70 (0.7) | 9 (0.3) | 17 (0.4) | 22 (1.8) | 22 (1.6) | |
| 0.10 – 0.29 | 333 (3.5) | 46 (1.5) | 155 (4.0) | 43 (3.4) | 89 (6.4) | |
| 0.30 – 0.49 | 931 (9.7) | 190 (6.2) | 502 (12.8) | 82 (6.5) | 157 (11.3) | |
| 0.50 – 0.79 | 2447 (25.5) | 708 (23.2) | 1124 (28.7) | 239 (19.0) | 376 (27.1) | |
| 0.80 – 1.49 | 4175 (43.4) | 1530 (50.0) | 1573 (40.2) | 546 (43.4) | 526 (37.9) | |
| 1.50 – 2.99 | 1480 (15.4) | 524 (17.1) | 472 (12.1) | 290 (23.1) | 194 (14.0) | |
| > 3.0 | 176 (1.8) | 50 (1.6) | 68 (1.7) | 35 (2.8) | 23 (1.7) | |
| Hemoglobin (mmol/L) | | | | | | <0.001 |
| Mean, (SD) | NA | 8.33 (1.27) | 7.97 (1.27) | NA | NA | |
| < 3.73 | 31 (0.3) | 5 (0.1) | 10 (0.2) | 4 (0.2) | 12 (0.7) | |
| 3.73 – 4.90 | 180 (1.6) | 46 (1.2) | 41 (0.9) | 29 (1.8) | 64 (3.8) | |
| 4.91 – 6.15 | 764 (6.6) | 178 (4.8) | 333 (7.4) | 90 (5.5) | 163 (9.6) | |
| 6.16 – 7.39 | 2101 (18.2) | 509 (13.8) | 971 (21.4) | 250 (15.3) | 371 (21.8) | |
| 7.4 – 9.25 | 6469 (56.0) | 2079 (56.3) | 2520 (55.7) | 941 (57.7) | 929 (54.5) | |
| >= 9.25 | 2010 (17.4) | 875 (23.7) | 653 (14.4) | 317 (19.4) | 165 (9.7) | |
| Platelets (x 10^9/L) | | | | | | <0.001 |
| Mean, (SD) | NA | 234 (104) | 221 (102) | NA | NA | |
| < 10 | 25 (0.2) | 4 (0.1) | 8 (0.2) | 6 (0.4) | 7 (0.4) | |
| 10 – 49 | 98 (0.9) | 19 (0.5) | 27 (0.6) | 28 (1.8) | 24 (1.4) | |
| 50 – 119 | 931 (8.4) | 201 (5.7) | 407 (9.5) | 123 (7.7) | 200 (12.0) | |
| 120 – 449 | 9681 (87.2) | 3177 (89.5) | 3695 (86.4) | 1411 (88.3) | 1398 (83.8) | |
| 450 – 799 | 350 (3.2) | 145 (4.1) | 135 (3.2) | 30 (1.9) | 40 (2.4) | |
| 800 – 1199 | 11 (0.1) | 5 (0.1) | 6 (0.1) | 0 (0.0) | 0 (0.0) | |
| Creatinine (µmol/L), Mean, (SD) | NA | 98 (107) | 116 (93) | NA | NA | NA |

CRP = C-Reactive Protein; IQR = Interquartile Range; SD = Standard Deviation

| Table S5: Baseline characteristics stratified by admission to a critical care unit | | | | |
|---|---|---|---|---|
| | **Overall** | **Ward** | **Critical care** | **P-value** |
| Total | 16511 | 10287 | 3916 | |
| **Age, years (%)** | | | | <0.001 |
| 18 – 25 | 239 (1.4) | 135 (1.3) | 36 (0.9) | |
| 26 – 35 | 704 (4.3) | 443 (4.3) | 90 (2.3) | |
| 36 – 45 | 1128 (6.8) | 657 (6.4) | 254 (6.5) | |
| 46 – 55 | 2299 (13.9) | 1328 (12.9) | 655 (16.7) | |
| 56 – 65 | 3280 (19.9) | 1792 (17.4) | 1089 (27.8) | |
| 66 – 75 | 3485 (21.1) | 2005 (19.5) | 1112 (28.4) | |
| 76 – 85 | 3720 (22.5) | 2601 (25.3) | 598 (15.3) | |
| >85 | 1656 (10.0) | 1326 (12.9) | 82 (2.1) | |
| **Sex, female (%)** | 6627 (40.2) | 4438 (43.2) | 1112 (28.4) | <0.001 |
| **Ethnicity** | | | | <0.001 |
| Arab | 496 (3.5) | 335 (3.6) | 161 (5.0) | |
| Asian | 898 (6.3) | 588 (6.3) | 257 (7.9) | |
| Black | 384 (2.7) | 253 (2.7) | 92 (2.8) | |
| Latin-American | 21 (0.1) | 14 (0.2) | 7 (0.2) | |
| White | 12120 (84.5) | 7838 (84.2) | 2586 (80.0) | |
| Other | 416 (2.9) | 286 (3.1) | 130 (4.0) | |
| **BMI (kg/m$^2$)** | | | | <0.001 |
| Underweight (<18.5) | 239 (2.3) | 169 (2.7) | 32 (1.1) | |
| Normal weight (18.5 – 24.9) | 3091 (29.7) | 2044 (32.8) | 642 (22.3) | |
| Overweight (25.0 – 29.9) | 3854 (37.0) | 2239 (35.9) | 1163 (40.4) | |
| Obese (30.0 – 34.9) | 2069 (19.9) | 1162 (18.6) | 641 (22.3) | |
| Morbidly obese (> 34.9) | 1157 (11.1) | 620 (9.9) | 398 (13.8) | |
| **Cardiovascular risk factors** | | | | |
| Diabetes | 4031 (24.9) | 2541 (25.1) | 1022 (26.8) | 0.039 |
| Hypertension | 7975 (49.5) | 4923 (49.0) | 1888 (49.8) | 0.422 |
| Peripheral arterial disease | 654 (4.8) | 405 (4.9) | 149 (4.5) | 0.426 |
| **Cardiac disease** | | | | |
| Any history of cardiac disease | 5198 (31.5) | 3519 (34.2) | 1036 (26.5) | <0.001 |
| Arrhythmia/conduction disorder | 2503 (15.3) | 1695 (16.5) | 452 (11.7) | <0.001 |
| Atrial fibrillation/flutter | 2004 (12.3) | 1309 (12.8) | 369 (9.6) | <0.001 |
| Coronary artery disease | 2420 (14.8) | 1504 (14.7) | 566 (14.6) | 0.958 |
| Myocardial infarction | 769 (4.8) | 463 (4.6) | 171 (4.5) | 0.801 |
| Heart failure | 1314 (8.1) | 876 (8.6) | 231 (6.0) | <0.001 |
| NYHA I/II | 324 (2.0) | 208 (2.0) | 46 (1.2) | 0.001 |
| NYHA III/IV | 246 (1.5) | 139 (1.4) | 58 (1.5) | 0.609 |
| Valvular heart disease | 579 (3.6) | 442 (4.3) | 82 (2.1) | <0.001 |
| **Comorbidities** | | | | |
| Chronic kidney disease | 2196 (13.5) | 1407 (13.9) | 440 (11.5) | <0.001 |
| COPD | 1563 (9.6) | 1096 (10.8) | 336 (8.8) | <0.001 |
| **Use cardiovascular drugs** | | | | |
| ACE-inhibitors | 3021 (18.8) | 1834 (18.0) | 697 (19.1) | 0.152 |
| Aldosterone antagonist | 563 (3.5) | 356 (3.5) | 103 (2.7) | 0.036 |
| Anti-platelet | 3044 (18.7) | 1958 (19.1) | 661 (17.6) | 0.042 |
| Angiotensin receptor blocker | 2159 (13.5) | 1295 (12.7) | 515 (14.1) | 0.033 |
| Calcium-blocker | 2769 (17.0) | 1629 (15.9) | 731 (19.5) | <0.001 |
| Diuretic | 3338 (20.5) | 2143 (20.9) | 686 (18.3) | 0.001 |
| Insulin | 1204 (7.4) | 730 (7.1) | 315 (8.2) | 0.034 |
| Lipid-lowering | 4789 (30.5) | 3300 (33.1) | 1015 (28.3) | <0.001 |
| Oral anti-diabetic | 2139 (13.0) | 1386 (13.5) | 512 (13.1) | 0.573 |

ACE = Angiotensin Converting Enzyme; BMI = Body Mass Index; COPD = Chronic Obstructive Pulmonary Disease; NYHA = New York Heart Association

| Table S6: Complaints, vitals and laboratory values at admission stratified by admission to a critical care unit | | | | |
|---|---|---|---|---|
| | **Overall** | **Ward** | **Critical care** | **P-value** |
| Total | 16511 | 10287 | 3916 | |
| **Admission** | | | | |
| Onset of symptoms to admission, days, median, [IQR] | 6 [2 – 9] | 5 [1 – 9] | 7 [3 – 9] | <0.001 |
| **Complaints at admission** | | | | |
| Anosmia | 723 (4.6) | 479 (4.9) | 122 (3.3) | <0.001 |
| Cough | 8158 (51.8) | 5303 (54.5) | 1957 (52.2) | 0.019 |
| Dyspnea | 7798 (49.8) | 4899 (50.7) | 2199 (59.1) | <0.001 |
| Fatigue | 4214 (26.8) | 2946 (30.3) | 922 (24.6) | <0.001 |
| Fever | 8638 (54.9) | 5565 (57.2) | 2188 (58.4) | 0.215 |
| Gastrointestinal symptoms | 3030 (19.2) | 2124 (21.8) | 702 (18.7) | <0.001 |
| Palpitations | 162 (1.0) | 106 (1.1) | 40 (1.1) | 0.986 |
| Sore throat | 1108 (7.0) | 734 (7.5) | 233 (6.2) | 0.008 |
| **Vitals at admission** | | | | |
| Temperature (°C) | | | | <0.001 |
| < 35.1 | 72 (0.6) | 45 (0.6) | 22 (0.8) | |
| 35.1 – 37.2 | 4898 (41.7) | 3417 (43.4) | 905 (33.7) | |
| 37.3 – 37.9 | 2546 (21.7) | 1754 (22.3) | 554 (20.6) | |
| 38.0 – 38.9 | 2920 (24.9) | 1856 (23.6) | 801 (29.8) | |
| 39.0 – 39.9 | 1139 (9.7) | 721 (9.2) | 338 (12.6) | |
| > 39.9 | 158 (1.3) | 79 (1.0) | 65 (2.4) | |
| Respiratory rate (breaths/min) | | | | <0.001 |
| < 16 | 1168 (10.8) | 764 (10.3) | 217 (8.6) | |
| 16 – 21 | 4914 (45.4) | 3634 (49.1) | 767 (30.5) | |
| 22 – 29 | 3219 (29.8) | 2171 (29.3) | 888 (35.4) | |
| > 29 | 1512 (14.0) | 832 (11.2) | 639 (25.4) | |
| Heart rate (beats/min) | | | | <0.001 |
| < 45 | 51 (0.4) | 33 (0.4) | 11 (0.4) | |
| 45 – 59 | 293 (2.5) | 214 (2.7) | 63 (2.3) | |
| 60 – 89 | 6037 (51.2) | 4106 (52.0) | 1223 (45.2) | |
| 90 – 119 | 4627 (39.3) | 3067 (38.8) | 1162 (42.9) | |
| > 119 | 775 (6.6) | 480 (6.1) | 248 (9.2) | |
| Systolic blood pressure (mmHg) | | | | 0.02 |
| < 80 | 79 (0.7) | 47 (0.6) | 21 (0.8) | |
| 80 – 99 | 566 (4.8) | 358 (4.5) | 153 (5.7) | |
| 100 – 119 | 2606 (22.1) | 1748 (22.1) | 622 (23.2) | |
| 120 – 139 | 4291 (36.5) | 2918 (36.9) | 923 (34.4) | |
| 140 – 179 | 3827 (32.5) | 2574 (32.6) | 860 (32.1) | |
| > 179 | 403 (3.4) | 258 (3.3) | 103 (3.8) | |
| Diastolic blood pressure (mmHg) | | | | <0.001 |
| < 40 | 44 (0.4) | 19 (0.2) | 23 (0.9) | |
| 40 – 59 | 1218 (10.4) | 732 (9.3) | 389 (14.5) | |
| 60 – 89 | 8452 (71.9) | 5716 (72.4) | 1866 (69.7) | |
| 90 – 109 | 1823 (15.5) | 1268 (16.1) | 355 (13.3) | |
| > 109 | 216 (1.8) | 157 (2.0) | 43 (1.6) | |
| Oxygen saturation (%) | | | | <0.001 |
| < 60 | 67 (0.6) | 20 (0.3) | 43 (1.6) | |
| 60 – 69 | 57 (0.5) | 16 (0.2) | 38 (1.4) | |
| 70 – 79 | 206 (1.8) | 74 (0.9) | 121 (4.4) | |
| 80 – 89 | 1313 (11.2) | 633 (8.0) | 585 (21.5) | |
| 90 – 95 | 4768 (40.6) | 3218 (40.9) | 1138 (41.7) | |
| 96 – 100 | 5330 (45.4) | 3904 (49.6) | 802 (29.4) | |
| **Laboratory values at admission** | | | | |
| CRP (mg/L) | | | | <0.001 |
| < 3 | 540 (4.7) | 310 (4.1) | 42 (1.6) | |
| 3 – 29 | 2664 (23.2) | 1844 (24.3) | 335 (12.4) | |
| 30 – 69 | 2605 (22.7) | 1945 (25.6) | 446 (16.5) | |
| 70 – 119 | 2225 (19.4) | 1561 (20.6) | 524 (19.4) | |
| 120 – 179 | 1657 (14.4) | 1045 (13.8) | 514 (19.0) | |
| 180 – 249 | 965 (8.4) | 539 (7.1) | 391 (14.5) | |

| | | | | |
|---|---|---|---|---|
| > 249 | 812 (7.1) | 347 (4.6) | 453 (16.7) | |
| White blood cell count (x 10^9/L) | | | | <0.001 |
| < 1.0 | 33 (0.3) | 22 (0.3) | 8 (0.3) | |
| 1.0 – 3.9 | 1327 (11.5) | 914 (12.0) | 248 (9.2) | |
| 4.0 – 7.9 | 6110 (53.0) | 4171 (54.8) | 1218 (44.9) | |
| 8.0 – 11.9 | 2773 (24.1) | 1793 (23.6) | 764 (28.2) | |
| 12.0 – 15.9 | 935 (8.1) | 515 (6.8) | 350 (12.9) | |
| 16.0 – 19.9 | 288 (2.5) | 175 (2.3) | 99 (3.7) | |
| > 20 | 60 (0.5) | 22 (0.3) | 23 (0.8) | |
| Lymphocyte count (x 10^9/L) | | | | <0.001 |
| < 0.1 | 70 (0.7) | 34 (0.5) | 21 (1.0) | |
| 0.10 – 0.29 | 333 (3.5) | 206 (3.1) | 94 (4.5) | |
| 0.30 – 0.49 | 931 (9.7) | 597 (9.0) | 263 (12.7) | |
| 0.50 – 0.79 | 2447 (25.5) | 1620 (24.5) | 626 (30.2) | |
| 0.80 – 1.49 | 4175 (43.4) | 2953 (44.7) | 839 (40.4) | |
| 1.50 – 2.99 | 1480 (15.4) | 1077 (16.3) | 196 (9.4) | |
| > 3.0 | 176 (1.8) | 116 (1.8) | 37 (1.8) | |
| Hemoglobin (mmol/L) | | | | 0.013 |
| < 3.73 | 31 (0.3) | 19 (0.2) | 5 (0.2) | |
| 3.73 – 4.90 | 180 (1.6) | 112 (1.5) | 47 (1.7) | |
| 4.91 – 6.15 | 764 (6.6) | 475 (6.2) | 222 (8.1) | |
| 6.16 – 7.39 | 2101 (18.2) | 1371 (18.0) | 499 (18.2) | |
| 7.4 – 9.25 | 6469 (56.0) | 4312 (56.5) | 1475 (53.9) | |
| >= 9.25 | 2010 (17.4) | 1348 (17.7) | 491 (17.9) | |
| Platelets (x 10^9/L) | | | | 0.073 |
| < 10 | 25 (0.2) | 10 (0.1) | 11 (0.4) | |
| 10 – 49 | 98 (0.9) | 58 (0.8) | 24 (0.9) | |
| 50 – 119 | 931 (8.4) | 595 (8.2) | 241 (9.1) | |
| 120 – 449 | 9681 (87.2) | 6386 (87.5) | 2275 (86.2) | |
| 450 – 799 | 350 (3.2) | 240 (3.3) | 86 (3.3) | |
| 800 – 1199 | 11 (0.1) | 8 (0.1) | 3 (0.1) | |

CRP = C-Reactive Protein; IQR = Interquartile Range

| Table S7: Baseline characteristics stratified by mortality | | | | |
|---|---|---|---|---|
| | **Overall** | **Discharged alive** | **Died during admission** | **P-value** |
| Total | 16511 | 13169 | 3342 | |
| **Age, years (%)** | | | | <0.001 |
| 18 – 25 | 239 (1.4) | 237 (1.8) | 2 (0.1) | |
| 26 – 35 | 704 (4.3) | 681 (5.2) | 23 (0.7) | |
| 36 – 45 | 1128 (6.8) | 1080 (8.2) | 48 (1.4) | |
| 46 – 55 | 2299 (13.9) | 2139 (16.2) | 160 (4.8) | |
| 56 – 65 | 3280 (19.9) | 2873 (21.8) | 407 (12.2) | |
| 66 – 75 | 3485 (21.1) | 2679 (20.3) | 806 (24.1) | |
| 76 – 85 | 3720 (22.5) | 2476 (18.8) | 1244 (37.2) | |
| >85 | 1656 (10.0) | 1004 (7.6) | 652 (19.5) | |
| **Sex, female (%)** | 6627 (40.2) | 5489 (41.7) | 1138 (34.1) | <0.001 |
| **Ethnicity** | | | | 0.001 |
| Arab | 496 (3.5) | 423 (3.7) | 73 (2.5) | |
| Asian | 898 (6.3) | 727 (6.4) | 171 (5.8) | |
| Black | 384 (2.7) | 322 (2.8) | 62 (2.1) | |
| Latin-American | 21 (0.1) | 18 (0.2) | 3 (0.1) | |
| White | 12120 (84.5) | 9545 (83.9) | 2575 (87.0) | |
| Other | 416 (2.9) | 339 (3.0) | 77 (2.6) | |
| **BMI (kg/m$^2$)** | | | | <0.001 |
| Underweight (<18.5) | 239 (2.3) | 168 (2.0) | 71 (3.5) | |
| Normal weight (18.5 – 24.9) | 3091 (29.7) | 2464 (29.4) | 627 (30.8) | |
| Overweight (25.0 – 29.9) | 3854 (37.0) | 3092 (36.9) | 762 (37.5) | |
| Obese (30.0 – 34.9) | 2069 (19.9) | 1706 (20.4) | 363 (17.9) | |
| Morbidly obese (> 34.9) | 1157 (11.1) | 947 (11.3) | 210 (10.3) | |
| **Cardiovascular risk factors** | | | | |
| Diabetes | 4031 (24.9) | 2966 (22.9) | 1065 (32.9) | <0.001 |
| Hypertension | 7975 (49.5) | 6013 (46.6) | 1962 (60.8) | <0.001 |
| Peripheral arterial disease | 654 (4.8) | 450 (4.0) | 204 (7.9) | <0.001 |
| **Cardiac disease** | | | | |
| Any history of cardiac disease | 5198 (31.5) | 3653 (27.7) | 1545 (46.2) | <0.001 |
| Arrhythmia/conduction disorder | 2503 (15.3) | 1714 (13.1) | 789 (23.9) | <0.001 |
| Atrial fibrillation/flutter | 2004 (12.3) | 1358 (10.4) | 646 (19.6) | <0.001 |
| Coronary artery disease | 2420 (14.8) | 1676 (12.8) | 744 (22.7) | <0.001 |
| Myocardial infarction | 769 (4.8) | 537 (4.1) | 232 (7.2) | <0.001 |
| Heart failure | 1314 (8.1) | 825 (6.3) | 489 (14.9) | <0.001 |
| NYHA I/II | 324 (2.0) | 221 (1.7) | 103 (3.1) | <0.001 |
| NYHA III/IV | 246 (1.5) | 139 (1.1) | 107 (3.2) | <0.001 |
| Valvular heart disease | 579 (3.6) | 388 (3.0) | 191 (5.8) | <0.001 |
| **Comorbidities** | | | | |
| Chronic kidney disease | 2196 (13.5) | 1460 (11.2) | 736 (22.6) | <0.001 |
| COPD | 1563 (9.6) | 1044 (8.1) | 519 (16.0) | <0.001 |
| **Use cardiovascular drugs** | | | | |
| ACE-inhibitors | 3021 (18.8) | 2318 (18.0) | 703 (22.2) | <0.001 |
| Aldosterone antagonist | 563 (3.5) | 412 (3.2) | 151 (4.7) | <0.001 |
| Anti-platelet | 3044 (18.7) | 2230 (17.1) | 814 (25.1) | <0.001 |
| Angiotensin receptor blocker | 2159 (13.5) | 1719 (13.4) | 440 (13.9) | 0.465 |
| Calcium-blocker | 2769 (17.0) | 2140 (16.4) | 629 (19.5) | <0.001 |
| Diuretic | 3338 (20.5) | 2351 (18.1) | 987 (30.5) | <0.001 |
| Insulin | 1204 (7.4) | 879 (6.7) | 325 (9.9) | <0.001 |
| Lipid-lowering | 4789 (30.5) | 3587 (28.5) | 1202 (38.6) | <0.001 |
| Oral anti-diabetic | 2139 (13.0) | 1609 (12.2) | 530 (15.9) | <0.001 |

ACE = Angiotensin Converting Enzyme; BMI = Body Mass Index; COPD = Chronic Obstructive Pulmonary Disease; NYHA = New York Heart Association

| Table S8: Complaints, vitals and laboratory values at admission stratified by mortality | | | | |
|---|---|---|---|---|
| | **Overall** | **Discharged alive** | **Died during admission** | **P-value** |
| Total | 16511 | 13169 | 3342 | |
| **Admission** | | | | |
| Onset of symptoms to admission, days, median, [IQR] | 6 [2 – 9] | 6 [2 – 10] | 4 [1 – 7] | <0.001 |
| **Complaints at admission** | | | | |
| Anosmia | 723 (4.6) | 666 (5.3) | 57 (1.8) | <0.001 |
| Cough | 8158 (51.8) | 6660 (52.9) | 1498 (47.5) | <0.001 |
| Dyspnea | 7798 (49.8) | 5996 (47.9) | 1802 (57.5) | <0.001 |
| Fatigue | 4214 (26.8) | 3345 (26.6) | 869 (27.5) | 0.288 |
| Fever | 8638 (54.9) | 6944 (55.2) | 1694 (53.7) | 0.135 |
| Gastrointestinal symptoms | 3030 (19.2) | 2494 (19.8) | 536 (17.0) | <0.001 |
| Palpitations | 162 (1.0) | 131 (1.0) | 31 (1.0) | 0.847 |
| Sore throat | 1108 (7.0) | 943 (7.5) | 165 (5.2) | <0.001 |
| **Vitals at admission** | | | | |
| Temperature (°C) | | | | <0.001 |
| < 35.1 | 72 (0.6) | 42 (0.4) | 30 (1.3) | |
| 35.1 – 37.2 | 4898 (41.7) | 3980 (42.4) | 918 (39.1) | |
| 37.3 – 37.9 | 2546 (21.7) | 2049 (21.8) | 497 (21.1) | |
| 38.0 – 38.9 | 2920 (24.9) | 2300 (24.5) | 620 (26.4) | |
| 39.0 – 39.9 | 1139 (9.7) | 891 (9.5) | 248 (10.6) | |
| > 39.9 | 158 (1.3) | 121 (1.3) | 37 (1.6) | |
| Respiratory rate (breaths/min) | | | | <0.001 |
| < 16 | 1168 (10.8) | 1014 (11.8) | 154 (7.0) | |
| 16 – 21 | 4914 (45.4) | 4165 (48.4) | 749 (33.8) | |
| 22 – 29 | 3219 (29.8) | 2431 (28.3) | 788 (35.6) | |
| > 29 | 1512 (14.0) | 989 (11.5) | 523 (23.6) | |
| Heart rate (beats/min) | | | | <0.001 |
| < 45 | 51 (0.4) | 40 (0.4) | 11 (0.5) | |
| 45 – 59 | 293 (2.5) | 226 (2.4) | 67 (2.8) | |
| 60 – 89 | 6037 (51.2) | 4904 (52.1) | 1133 (48.0) | |
| 90 – 119 | 4627 (39.3) | 3728 (39.6) | 899 (38.1) | |
| > 119 | 775 (6.6) | 523 (5.6) | 252 (10.7) | |
| Systolic blood pressure (mmHg) | | | | <0.001 |
| < 80 | 79 (0.7) | 44 (0.5) | 35 (1.5) | |
| 80 – 99 | 566 (4.8) | 402 (4.3) | 164 (6.9) | |
| 100 – 119 | 2606 (22.1) | 2051 (21.8) | 555 (23.5) | |
| 120 – 139 | 4291 (36.5) | 3554 (37.8) | 737 (31.2) | |
| 140 – 179 | 3827 (32.5) | 3047 (32.4) | 780 (33.0) | |
| > 179 | 403 (3.4) | 310 (3.3) | 93 (3.9) | |
| Diastolic blood pressure (mmHg) | | | | <0.001 |
| < 40 | 44 (0.4) | 26 (0.3) | 18 (0.8) | |
| 40 – 59 | 1218 (10.4) | 832 (8.9) | 386 (16.4) | |
| 60 – 89 | 8452 (71.9) | 6816 (72.6) | 1636 (69.4) | |
| 90 – 109 | 1823 (15.5) | 1559 (16.6) | 264 (11.2) | |
| > 109 | 216 (1.8) | 161 (1.7) | 55 (2.3) | |
| Oxygen saturation (%) | | | | <0.001 |
| < 60 | 67 (0.6) | 31 (0.3) | 36 (1.5) | |
| 60 – 69 | 57 (0.5) | 23 (0.2) | 34 (1.4) | |
| 70 – 79 | 206 (1.8) | 95 (1.0) | 111 (4.7) | |
| 80 – 89 | 1313 (11.2) | 861 (9.2) | 452 (19.1) | |
| 90 – 95 | 4768 (40.6) | 3800 (40.5) | 968 (40.9) | |
| 96 – 100 | 5330 (45.4) | 4563 (48.7) | 767 (32.4) | |
| **Laboratory values at admission** | | | | |
| CRP (mg/L) | | | | <0.001 |
| < 3 | 540 (4.7) | 512 (5.6) | 28 (1.2) | |
| 3 – 29 | 2664 (23.2) | 2363 (25.8) | 301 (13.0) | |
| 30 – 69 | 2605 (22.7) | 2146 (23.5) | 459 (19.8) | |
| 70 – 119 | 2225 (19.4) | 1734 (19.0) | 491 (21.1) | |
| 120 – 179 | 1657 (14.4) | 1215 (13.3) | 442 (19.0) | |

| | | | | |
|---|---|---|---|---|
| 180 – 249 | 965 (8.4) | 662 (7.2) | 303 (13.0) | |
| > 249 | 812 (7.1) | 513 (5.6) | 299 (12.9) | |
| White blood cell count (x 10^9/L) | | | | <0.001 |
| < 1.0 | 33 (0.3) | 24 (0.3) | 9 (0.4) | |
| 1.0 – 3.9 | 1327 (11.5) | 1106 (12.0) | 221 (9.5) | |
| 4.0 – 7.9 | 6110 (53.0) | 5034 (54.7) | 1076 (46.5) | |
| 8.0 – 11.9 | 2773 (24.1) | 2181 (23.7) | 592 (25.6) | |
| 12.0 – 15.9 | 935 (8.1) | 643 (7.0) | 292 (12.6) | |
| 16.0 – 19.9 | 288 (2.5) | 186 (2.0) | 102 (4.4) | |
| > 20 | 60 (0.5) | 37 (0.4) | 23 (1.0) | |
| Lymphocyte count (x 10^9/L) | | | | <0.001 |
| < 0.1 | 70 (0.7) | 46 (0.6) | 24 (1.2) | |
| 0.10 – 0.29 | 333 (3.5) | 204 (2.7) | 129 (6.6) | |
| 0.30 – 0.49 | 931 (9.7) | 638 (8.3) | 293 (15.0) | |
| 0.50 – 0.79 | 2447 (25.5) | 1851 (24.2) | 596 (30.5) | |
| 0.80 – 1.49 | 4175 (43.4) | 3486 (45.5) | 689 (35.2) | |
| 1.50 – 2.99 | 1480 (15.4) | 1290 (16.8) | 190 (9.7) | |
| > 3.0 | 176 (1.8) | 142 (1.9) | 34 (1.7) | |
| Hemoglobin (mmol/L) | | | | <0.001 |
| < 3.73 | 31 (0.3) | 17 (0.2) | 14 (0.6) | |
| 3.73 – 4.90 | 180 (1.6) | 137 (1.5) | 43 (1.8) | |
| 4.91 – 6.15 | 764 (6.6) | 526 (5.7) | 238 (10.2) | |
| 6.16 – 7.39 | 2101 (18.2) | 1584 (17.2) | 517 (22.1) | |
| 7.4 – 9.25 | 6469 (56.0) | 5275 (57.2) | 1194 (51.0) | |
| >= 9.25 | 2010 (17.4) | 1677 (18.2) | 333 (14.2) | |
| Platelets (x 10^9/L) | | | | <0.001 |
| < 10 | 25 (0.2) | 15 (0.2) | 10 (0.4) | |
| 10 – 49 | 98 (0.9) | 75 (0.8) | 23 (1.0) | |
| 50 – 119 | 931 (8.4) | 651 (7.3) | 280 (12.5) | |
| 120 – 449 | 9681 (87.2) | 7827 (88.3) | 1854 (83.0) | |
| 450 – 799 | 350 (3.2) | 286 (3.2) | 64 (2.9) | |
| 800 – 1199 | 11 (0.1) | 9 (0.1) | 2 (0.1) | |

CRP = C-Reactive Protein; IQR = Interquartile Range

| **Table S9: Outcome at discharge stratified by data source and age** | | | | | | |
|---|---|---|---|---|---|---|
| | **Overall** | **CAPACITY-COVID** | | **LEOSS** | | **P-value** |
| | | **≤65 years** | **>65 years** | **≤65 years** | **>65 years** | |
| Total | 16511 | 4303 | 5501 | 3347 | 3360 | |
| **Admission** | | | | | | |
| Duration of hospitalization, days, median [IQR] | NA | 8 [4 – 18] | 9 [5 – 18] | NA | NA | NA |
| Admission to a critical care unit | 3916 (27.6) | 1314 (30.5) | 954 (17.3) | 810 (37.2) | 838 (37.7) | <0.001 |
| Duration of stay critical care unit, days, median [IQR] | NA | 12 [6 – 23] | 12 [6 – 23] | NA | NA | NA |
| **Treatment** | | | | | | |
| Invasive ventilation | 2680 (16.2) | 1033 (24.1) | 763 (13.9) | 440 (13.2) | 444 (13.2) | <0.001 |
| Non-invasive ventilation | 1818 (11.1) | 723 (17.1) | 526 (9.6) | 284 (8.5) | 285 (8.5) | <0.001 |
| ECMO | 316 (1.9) | 106 (2.5) | 16 (0.3) | 145 (4.3) | 49 (1.5) | <0.001 |
| **Complications** | | | | | | |
| Cardiac | | | | | | |
| Endocarditis | NA | 2 (0.0) | 7 (0.1) | NA | NA | NA |
| Heart failure de novo | 197 (1.2) | 30 (0.7) | 101 (1.8) | 28 (0.8) | 38 (1.1) | <0.001 |
| Myocardial infarction | 95 (0.6) | 14 (0.3) | 32 (0.6) | 11 (0.3) | 38 (1.1) | <0.001 |
| Myocarditis | 37 (0.2) | 13 (0.3) | 14 (0.3) | 8 (0.2) | 2 (0.1) | 0.140 |
| Pericarditis | NA | 13 (0.3) | 2 (0.0) | NA | NA | NA |
| Ventricular arrhythmia | NA | 17 (0.4) | 34 (0.6) | NA | NA | NA |
| Thromboembolic | | | | | | |
| Pulmonary embolism | 569 (3.5) | 238 (5.6) | 218 (4.0) | 56 (1.7) | 57 (1.7) | <0.001 |
| Stroke | 106 (0.6) | 21 (0.5) | 50 (0.9) | 5 (0.2) | 30 (0.9) | <0.001 |
| Venous thrombosis | 121 (0.7) | 29 (0.7) | 20 (0.4) | 37 (1.1) | 35 (1.1) | <0.001 |
| **Outcome** | | | | | | |
| Deceased | 3342 (20.2) | 422 (9.8) | 1819 (33.1) | 218 (6.5) | 883 (26.3) | <0.001 |

ECMO = Extracorporeal Membrane Oxygenation; IQR = Interquartile Range

| Table S10: Outcome at discharge stratified by admission to a critical care unit | | | | |
|---|---|---|---|---|
| | **Overall** | **Ward** | **Critical care** | **P-value** |
| Total | 16511 | 10287 | 3916 | |
| **Admission** | | | | |
| Duration of hospitalization, days, median [IQR] | 9 [5 – 18] | 7 [4 – 13] | 21 [12 – 35] | <0.001 |
| **Treatment** | | | | |
| Invasive ventilation | 2680 (16.2) | 0 (0.0) | 2680 (68.7) | <0.001 |
| Non-invasive ventilation | 1818 (11.1) | 35 (0.3) | 1783 (46.9) | <0.001 |
| ECMO | 316 (1.9) | 0 (0.0) | 316 (8.2) | <0.001 |
| **Complications** | | | | |
| Cardiac | | | | |
| Myocarditis | 37 (0.2) | 13 (0.1) | 23 (0.6) | <0.001 |
| Myocardial infarction | 95 (0.6) | 31 (0.3) | 61 (1.6) | <0.001 |
| Heart failure de novo | 197 (1.2) | 97 (0.9) | 93 (2.4) | <0.001 |
| Thromboembolic | | | | |
| Pulmonary embolism | 569 (3.5) | 144 (1.4) | 405 (10.5) | <0.001 |
| Stroke | 106 (0.6) | 52 (0.5) | 48 (1.2) | <0.001 |
| Venous thrombosis | 121 (0.7) | 28 (0.3) | 86 (2.2) | <0.001 |
| **Outcome** | | | | |
| Deceased | 3342 (20.2) | 1799 (17.5) | 1317 (33.6) | <0.001 |

ECMO = Extracorporeal Membrane Oxygenation; IQR = Interquartile Range

| Table S11: Outcome at discharge stratified by mortality | | | | |
|---|---|---|---|---|
| | **Overall** | **Discharged alive** | **Died during admission** | **P-value** |
| Total | 16511 | 13169 | 3342 | |
| **Admission** | | | | |
| Duration of hospitalization, days, median [IQR] | 9 [5 – 18] | 9 [5 – 18] | 8 [5 – 16] | 0.002 |
| Admission to a critical care unit | 3916 (27.6) | 2599 (23.4) | 1317 (42.3) | <0.001 |
| Duration of stay critical care unit, days, median [IQR] | 12 [6 – 23] | 13 [6 – 25] | 11 [6 – 19] | <0.001 |
| **Treatment** | | | | |
| Invasive ventilation | 2680 (16.2) | 1608 (12.2) | 1072 (32.1) | <0.001 |
| Non-invasive ventilation | 1818 (11.1) | 1267 (9.7) | 551 (16.7) | <0.001 |
| ECMO | 316 (1.9) | 148 (1.1) | 168 (5.1) | <0.001 |
| **Complications** | | | | |
| Cardiac | | | | |
| Myocarditis | 37 (0.2) | 24 (0.2) | 13 (0.4) | 0.039 |
| Myocardial infarction | 95 (0.6) | 40 (0.3) | 55 (1.7) | <0.001 |
| Heart failure de novo | 197 (1.2) | 92 (0.7) | 105 (3.1) | <0.001 |
| Thromboembolic | | | | |
| Pulmonary embolism | 569 (3.5) | 404 (3.1) | 165 (5.0) | <0.001 |
| Stroke | 106 (0.6) | 63 (0.5) | 43 (1.3) | <0.001 |
| Venous thrombosis | 121 (0.7) | 86 (0.7) | 35 (1.1) | 0.021 |

ECMO = Extracorporeal Membrane Oxygenation; IQR = Interquartile Range

| **Table S12: Subgroup analyses of having any history of cardiac disease among clinically relevant subgroups** | | | |
|---|---|---|---|
| | **p-value interaction** | **RR [95% CI]** | **p-value subgroup** |
| **Multivariable adjusted*** | | | |
| **Age** | 0.10 | | |
| ≤65 years | | 1.25 [1.04 – 1.50] | 0.02 |
| >65 years | | 1.06 [0.99 – 1.13] | 0.08 |
| **Sex** | 0.09 | | |
| Female | | 1.16 [1.05 – 1.29] | <0.001 |
| Male | | 1.04 [0.97 – 1.12] | 0.25 |
| **BMI (kg/m$^2$)** | 0.18 | | |
| <30 | | 1.11 [1.03 – 1.20] | 0.01 |
| ≥30 | | 1.00 [0.87 -1.14] | 0.98 |
| **Diabetes** | 0.27 | | |
| No | | 1.12 [1.03 – 1.20] | <0.001 |
| Yes | | 1.03 [0.93 – 1.14] | 0.52 |
| **Hypertension** | 0.21 | | |
| No | | 1.14 [1.03 – 1.26] | 0.01 |
| Yes | | 1.05 [0.98 – 1.14] | 0.18 |
| **Chronic kidney disease** | 0.33 | | |
| No | | 1.10 [1.02 – 1.18] | 0.01 |
| Yes | | 1.03 [0.91 – 1.16] | 0.69 |
| **COPD** | 0.54 | | |
| No | | 1.07 [1.00 – 1.15] | 0.04 |
| Yes | | 1.13 [0.98 – 1.30] | 0.10 |

* Multivariable models are adjusted for the following covariates: age, sex, BMI, hypertension, CKD, COPD, diabetes and region of inclusion (Central Europe, Middle – East, Netherlands/Belgium, Russia, Southern – Europe and the United Kingdom).

| Table S13: Sensitivity analyses between centers with and without a selective inclusion strategy | | | | |
|---|---|---|---|---|
| | **Total cohort** | | **Cohort with exclusion of patients included from centers with selective inclusion** | |
| | **RR [95% CI]** | **p-value*** | **RR [95% CI]** | **p-value*** |
| **Crude** | | | | |
| Any cardiac history | 1.87 [1.76 – 1.99] | <0.01 | 1.89 [1.78 – 2.01] | <0.01 |
| Arrhythmia/conduction disorder | 1.73 [1.62 – 1.85] | <0.01 | 1.75 [1.63 – 1.88] | <0.01 |
| Atrial fibrillation | 1.73 [1.61 – 1.86] | <0.01 | 1.76 [1.63 – 1.89] | <0.01 |
| Coronary artery disease | 1.68 [1.57 – 1.80] | <0.01 | 1.70 [1.58 – 1.83] | <0.01 |
| Myocardial infarction | 1.55 [1.39 – 1.74] | <0.01 | 1.56 [1.39 – 1.75] | <0.01 |
| Heart failure | 1.99 [1.84 – 2.15] | <0.01 | 2.00 [1.84 – 2.16] | <0.01 |
| NYHA I/II | 1.72 [1.47 – 2.02] | <0.01 | 1.72 [1.46 – 2.02] | <0.01 |
| NYHA III/IV | 2.37 [2.06 – 2.74] | <0.01 | 2.39 [2.07 – 2.76] | <0.01 |
| Valvular disease | 1.68 [1.49 – 1.89] | <0.01 | 1.68 [1.47 – 1.91] | <0.01 |
| **Age and sex adjusted** | | | | |
| Any cardiac history | 1.13 [1.06 – 1.20] | <0.018 | 1.13 [1.07 – 1.21] | <0.018 |
| Arrhythmia/conduction disorder | 1.06 [0.99 – 1.13] | 0.72 | 1.06 [0.99 – 1.14] | 0.90 |
| Atrial fibrillation | 1.07 [1.00 – 1.15] | 0.70 | 1.08 [1.00 – 1.16] | 0.50 |
| Coronary artery disease | 1.14 [1.06 – 1.22] | <0.018 | 1.14 [1.06 – 1.22] | <0.018 |
| Myocardial infarction | 1.10 [0.99 – 1.22] | 0.72 | 1.09 [0.98 – 1.22] | 0.96 |
| Heart failure | 1.31 [1.21 – 1.42] | <0.018 | 1.30 [1.20 – 1.41] | <0.018 |
| NYHA I/II | 1.16 [0.98 – 1.36] | 0.72 | 1.14 [0.97 – 1.35] | 0.96 |
| NYHA III/IV | 1.53 [1.32 – 1.78] | <0.018 | 1.53 [1.31 – 1.77] | <0.018 |
| Valvular disease | 1.11 [0.98 – 1.25] | 0.72 | 1.10 [0.97 – 1.25] | 0.96 |
| **Multivariable adjusted**** | | | | |
| Any cardiac disease | 1.08 [1.02 – 1.15] | 0.12 | 1.09 [1.02 – 1.17] | 0.12 |
| Arrhythmia/conduction disorder | 1.01 [0.94 – 1.08] | 1.00 | 1.02 [0.95 – 1.10] | 1.00 |
| Atrial fibrillation | 1.04 [0.97 – 1.12] | 1.00 | 1.05 [0.97 – 1.14] | 0.96 |
| Coronary artery disease | 1.10 [1.03 – 1.18] | 0.12 | 1.11 [1.03 – 1.19] | 0.12 |
| Myocardial infarction | 1.07 [0.96 – 1.19] | 1.00 | 1.07 [0.95 – 1.19] | 1.00 |
| Heart failure | 1.19 [1.10 – 1.30] | <0.018 | 1.19 [1.10 – 1.30] | <0.018 |
| NYHA I/II | 1.05 [0.90 – 1.24] | 1.00 | 1.04 [0.89 – 1.23] | 1.00 |
| NYHA III/IV | 1.41 [1.20 – 1.64] | <0.018 | 1.41 [1.21 – 1.65] | <0.018 |
| Valvular disease | 1.03 [0.91 – 1.16] | 1.00 | 1.02 [0.90 – 1.16] | 1.00 |

* p-values are adjusted for multiple testing by the Holm-Bonferroni method.

** Multivariable models are adjusted for the following covariates: age, sex, BMI, hypertension, CKD, COPD, diabetes and region of inclusion (Central Europe, Middle – East, Netherlands/Belgium, Russia, Southern – Europe and the United Kingdom). The models testing the association between a specific heart disease subtype and in-hospital mortality are also adjusted for the presence of other cardiac comorbidities (i.e. the model testing the association between heart failure and in-hospital mortality is adjusted for arrhythmia/conduction disorder, coronary artery disease and valvular disease).

| Table S14: Associations across data sources between pre-existent heart disease subtypes and in-hospital mortality | | | | | | |
|---|---|---|---|---|---|---|
| | CAPACITY-COVID | | LEOSS | | Total | |
| | RR [95% CI] | p-value* | RR [95% CI] | p-value* | RR [95% CI] | p-value* |
| **Crude** | | | | | | |
| Any cardiac history | 1.60 [1.49 – 1.72] | <0.01 | 2.43 [2.19 – 2.70] | <0.01 | 1.87 [1.76 – 1.99] | <0.01 |
| Arrhythmia/conduction disorder | 1.57 [1.44 – 1.70] | <0.01 | 2.10 [1.87 – 2.35] | <0.01 | 1.73 [1.62 – 1.85] | <0.01 |
| Atrial fibrillation | 1.61 [1.47 – 1.76] | <0.01 | 2.09 [1.86 – 2.35] | <0.01 | 1.73 [1.61 – 1.86] | <0.01 |
| Coronary artery disease | 1.49 [1.37 – 1.63] | <0.01 | 2.14 [1.91 – 2.39] | <0.01 | 1.68 [1.57 – 1.80] | <0.01 |
| Myocardial infarction | 1.35 [1.16 – 1.58] | <0.01 | 1.98 [1.70 – 2.32] | <0.01 | 1.55 [1.39 – 1.74] | <0.01 |
| Heart failure | 1.76 [1.59 – 1.94] | <0.01 | 2.52 [2.23 – 2.85] | <0.01 | 1.99 [1.84 – 2.15] | <0.01 |
| NYHA I/II | 1.58 [1.34 – 1.87] | <0.01 | 2.02 [1.55 – 2.63] | <0.01 | 1.72 [1.47 – 2.02] | <0.01 |
| NYHA III/IV | 2.17 [1.76 – 2.67] | <0.01 | 2.94 [2.46 – 3.52] | <0.01 | 2.37 [2.06 – 2.74] | <0.01 |
| Valvular disease | 1.54 [1.34 – 1.76] | <0.01 | 1.87 [1.46 – 2.40] | <0.01 | 1.68 [1.49 – 1.89] | <0.01 |
| **Age and sex adjusted** | | | | | | |
| Any cardiac history | 1.04 [0.97 – 1.12] | 1.00 | 1.30 [1.17 – 1.46] | <0.018 | 1.13 [1.06 – 1.20] | <0.018 |
| Arrhythmia/conduction disorder | 1.03 [0.95 – 1.12] | 1.00 | 1.14 [1.01 – 1.27] | 0.24 | 1.06 [0.99 – 1.13] | 0.72 |
| Atrial fibrillation | 1.06 [0.97 – 1.16] | 1.00 | 1.15 [1.03 – 1.29] | 0.20 | 1.07 [1.00 – 1.15] | 0.70 |
| Coronary artery disease | 1.07 [0.98 – 1.17] | 1.00 | 1.30 [1.16 – 1.46] | <0.018 | 1.14 [1.06 – 1.22] | <0.018 |
| Myocardial infarction | 1.0 [0.86 – 1.16] | 1.00 | 1.29 [1.10 – 1.51] | <0.018 | 1.10 [0.99 – 1.22] | 0.72 |
| Heart failure | 1.22 [1.11 – 1.35] | <0.018 | 1.51 [1.33 – 1.71] | <0.018 | 1.31 [1.21 – 1.42] | <0.018 |
| NYHA I/II | 1.11 [0.94 – 1.32] | 1.00 | 1.24 [0.95 – 1.63] | 0.91 | 1.16 [0.98 – 1.36] | 0.72 |
| NYHA III/IV | 1.48 [1.19 – 1.84] | <0.018 | 1.72 [1.42 – 2.08] | <0.018 | 1.53 [1.32 – 1.78] | <0.018 |
| Valvular disease | 1.08 [0.95 – 1.24] | 1.00 | 1.05 [0.83 – 1.34] | 1.00 | 1.11 [0.98 – 1.25] | 0.72 |
| **Multivariable adjusted**** | | | | | | |
| Any cardiac disease | 1.03 [0.96 – 1.11] | 1.00 | 1.18 [1.05 – 1.32] | <0.018 | 1.08 [1.02 – 1.15] | 0.12 |
| Arrhythmia/conduction disorder | 1.01 [0.93 – 1.11] | 1.00 | 0.99 [0.88 – 1.12] | 1.00 | 1.01 [0.94 – 1.08] | 1.00 |
| Atrial fibrillation | 1.04 [0.95 – 1.15] | 1.00 | 1.01 [0.90 – 1.14] | 1.00 | 1.04 [0.97 – 1.12] | 1.00 |
| Coronary artery disease | 1.05 [0.96 – 1.15] | 1.00 | 1.16 [1.03 – 1.30] | 0.20 | 1.10 [1.03 – 1.18] | 0.12 |
| Myocardial infarction | 0.98 [0.84 – 1.13] | 1.00 | 1.13 [0.96 – 1.32] | 0.91 | 1.07 [0.96 – 1.19] | 1.00 |
| Heart failure | 1.11 [1.00 – 1.24] | 0.75 | 1.30 [1.13 – 1.49] | <0.018 | 1.19 [1.10 – 1.30] | <0.018 |
| NYHA I/II | 1.02 [0.86 – 1.22] | 1.00 | 1.11 [0.85 – 1.46] | 1.00 | 1.05 [0.90 – 1.24] | 1.00 |
| NYHA III/IV | 1.33 [1.06 – 1.66] | 0.32 | 1.45 [1.19 – 1.77] | <0.018 | 1.41 [1.20 – 1.64] | <0.018 |
| Valvular disease | 1.06 [0.93 – 1.22] | 1.00 | 0.94 [0.74 – 1.20] | 1.00 | 1.03 [0.91 – 1.16] | 1.00 |

* p-values are adjusted for multiple testing by the Holm-Bonferroni method.

** Multivariable models are adjusted for the following covariates: age, sex, BMI, hypertension, CKD, COPD, diabetes and region of inclusion (Central Europe, Middle – East, Netherlands/Belgium, Russia, Southern – Europe and the United Kingdom). The models testing the association between a specific heart disease subtype and in-hospital mortality are also adjusted for the presence of other cardiac comorbidities (i.e. the model testing the association between heart failure and in-hospital mortality is adjusted for arrhythmia/conduction disorder, coronary artery disease and valvular disease).

| Table S15: Subgroup analyses for patients that were admitted to a critical care unit | | | | | |
|---|---|---|---|---|---|
| | | Patients only admitted to the ward | | Patients admitted to a critical care unit | |
| | p-value interaction | RR [95% CI] | p-value subgroup | RR [95% CI] | p-value subgroup |
| **Multivariable adjusted**** | | | | | |
| Any cardiac disease | <0.001 | 1.26 [1.16 – 1.37] | <0.001 | 0.96 [0.88 – 1.06] | 0.40 |
| Arrhythmia/conduction disorder | <0.001 | 1.17 [1.07 – 1.28] | <0.001 | 0.82 [0.73 – 0.92] | <0.001 |
| Atrial fibrillation | <0.001 | 1.22 [1.11 – 1.33] | <0.001 | 0.80 [0.71 – 0.91] | <0.001 |
| Coronary artery disease | 0.07 | 1.18 [1.08 – 1.29] | 0.08 | 1.03 [0.93 – 1.15] | 0.55 |
| Myocardial infarction | 0.57 | 1.12 [0.97 – 1.29] | 0.12 | 1.05 [0.89 – 1.24] | 0.57 |
| Heart failure | 0.01 | 1.29 [1.17 – 1.43] | <0.001 | 1.00 [0.86 – 1.17] | 0.97 |
| NYHA I/II | 0.07 | 1.15 [0.98 – 1.35] | 0.10 | 0.91 [0.71 – 1.17] | 0.46 |
| NYHA III/IV | 0.01 | 1.55 [1.28 – 1.87] | <0.001 | 1.09 [0.88 – 1.35] | 0.41 |
| Valvular disease | <0.001 | 1.23 [1.07 – 1.41] | <0.001 | 0.72 [0.55 – 0.95] | 0.02 |

* Multivariable models are adjusted for the following covariates: age, sex, BMI, hypertension, CKD, COPD, diabetes and region of inclusion (Central Europe, Middle – East, Benelux, Russia, Southern – Europe and the United Kingdom). The models testing the association between a specific heart disease subtype and in-hospital mortality are also adjusted for the presence of other cardiac comorbidities (i.e. the model testing the association between heart failure and in-hospital mortality is adjusted for arrhythmia/conduction disorder, coronary artery disease and valvular disease).
